# Latent Brain-Symptom Dimensions Characterize Heterogeneity in Clinical High Risk for Psychosis

**DOI:** 10.64898/2026.10.08.26364664

**Authors:** Jiayu Feng, Sarah Genon, Simon B. Eickhoff, Fali Li, Lin Jiang, Yulin Wang, Debo Dong

**Affiliations:** Key Laboratory of Cognition and Personality, Ministry of Education, Faculty of Psychology, Southwest University, Chongqing, 400715, China; Institute of Neuroscience and Medicine, Brain and Behaviour (INM-7), Research Centre Jülich, Jülich, Germany; Institute for Systems Neuroscience, Medical Faculty, Heinrich-Heine University Düsseldorf, Düsseldorf, Germany; The Clinical Hospital of Chengdu Brain Science Institute, MOE Key Lab for Neuroinformation, School of Life Science and Technology, University of Electronic Science and Technology of China, Chengdu, 611731, China; Department of Radiology, Sichuan Provincial People’s Hospital, University of Electronic Science and Technology of China, ChengDu, 610072, China; Chinese Academy of Sciences Sichuan Translational Medicine Research Hospital, Chengdu, 610072, China; Brain-Computer Interface and Brain-Inspired Intelligence Key Laboratory of Sichuan Province, University of Electronic Science and Technology of China, Chengdu, 611731, China; Research Unit of Neuroinformation, Chinese Academy of Medical Sciences, 2019RU035, Chengdu, 611731, China; Sleep and NeuroImaging Center, Faculty of Psychology, Southwest University, Chongqing, 400715, China

**Keywords:** Clinical high risk, Psychosis, Resting-state functional connectivity, Heterogeneity, Dimension approach, Biotypes

## Abstract

**BACKGROUND:** Clinical high risk (CHR) for psychosis is characterized by substantial heterogeneity in symptoms, functioning, and outcomes. Although distributed resting-state functional connectivity (RSFC) abnormalities have been reported, it remains unclear whether this heterogeneity can be organized into consistent multivariate brain-symptom dimensions and clinically meaningful subgroups.

**METHODS:** We analyzed RSFC and 34 clinical variables from 701 CHR participants in the Accelerating Medicines Partnership® Schizophrenia Program (AMP® SCZ). Partial least squares analysis identified latent brain-symptom dimensions, which were further examined using transcriptomic annotation, independent psychosocial and cognitive measures, hierarchical clustering, and Month 2 follow-up data.

**RESULTS:** Two significant brain-symptom dimensions emerged. The first reflected general psychopathology and was associated with somatomotor-salience and higher-order connectivity, with enrichment for genes related to neuronal development. The second, capturing symptom differentiation, spanned negative/internalizing features with psychobehavioral overactivation and involved cerebellar-thalamic-somatomotor circuitry, with enrichment for genes related to synaptic signaling. These dimensions showed dissociable associations with broader psychosocial and cognitive measures, and their loading patterns were substantially reproducible at Month 2. Clustering based on these dimensions identified two biotypes with distinct clinical and connectivity profiles that remained broadly similar at Month 2 despite divergent short-term RSFC changes.

**CONCLUSIONS:** These findings support complementary dimensional and categorical representations of CHR heterogeneity that link clinical variation with distributed brain connectivity and molecular signatures, while demonstrating short-term reproducibility of the identified dimensions and divergent RSFC changes across biotypes. This multilevel framework may help characterize biologically meaningful variation in psychosis risk.

## Introduction

Clinical high risk (CHR) for psychosis represents a critical opportunity for early detection and preventive intervention, providing a unique opportunity to characterize pathophysiological processes before frank psychotic disorder onset (1,2). Yet CHR is not a unitary clinical state. Instead, it is marked by substantial heterogeneity in symptom expression, functional impairment, and longitudinal outcomes (3–7). While only a minority of CHR individuals transition to psychosis (8,9), many remain clinically stable or remit over time (3,10). This heterogeneity poses a challenge for understanding the neurobiological basis of the psychosis risk state and the multidimensional nature of clinical presentation in CHR (11,12).

Previous studies have documented broad abnormalities in CHR across multiple domains, including clinical symptoms (e.g., attenuated positive, negative, and depressive symptoms) (7), cognitive deficits (2,13), and functional impairments (14). At the neural level, resting-state functional connectivity (RSFC) studies have similarly revealed distributed alterations, with convergent evidence implicating large-scale systems such as the salience network (15). Together, these findings point to a multidimensional phenotype accompanied by distributed alterations in functional brain organization. Studies in CHR have primarily focused on associations between specific connectivity measures and individual clinical or cognitive features (15,16). Recent multivariate studies in psychosis and transdiagnostic populations have extended this perspective by jointly modeling distributed brain measures and multiple clinical features, identifying latent patterns that covary across individuals (17–19). However, it remains unclear whether such relationships can be characterized in CHR and account for meaningful variation in clinical profiles and brain architecture. Addressing this question requires an analytic framework capable of identifying coordinated many-to-many associations between high-dimensional RSFC patterns and interindividual variation in clinical symptoms (19,20). Partial least squares (PLS) is well suited to this goal, as it extracts latent components that maximize covariance between multivariate brain and symptom data (17,21), thereby providing a parsimonious account of brain-symptom organization in clinically heterogeneous populations (22).

Identifying latent brain-symptom dimensions, however, is only a first step. To further evaluate their biological relevance, these dimensions can be examined in relation to gene expression profiles, rather than remaining purely phenomenological (23,24). Transcriptomic approaches provide an opportunity to assess whether macroscale neuroimaging phenotypes correspond to spatial patterns of gene expression, offering a molecular annotation of large-scale functional organization (22,25–27). In parallel, it is important to examine whether the identified dimensions generalize beyond the discovery symptom set. In CHR, cognitive performance, social functioning, and related psychosocial outcomes are closely related to prognosis and represent important targets for early intervention (28–30). Dimensions capturing meaningful variation in the psychosis risk state are therefore expected to show dissociable associations with these broader domains, rather than being confined to core symptom measures alone. Such dimensions also provide a basis for categorical analyses (31–33). While dimensional analyses describe continuous brain-symptom patterns across individuals, clustering can organize individuals with similar patterns into more homogeneous subgroups with distinct clinical and neural profiles. However, prior studies proposing subgroups or biotypes within CHR (12,22,30) have rarely integrated multivariate models that jointly capture brain and symptom information. Integrating dimensional and categorical approaches may therefore provide a more comprehensive characterization of heterogeneity in CHR.

In the present study, we used brain imaging and comprehensive clinical data from the Accelerating Medicines Partnership® Schizophrenia Program (AMP® SCZ) (34) to examine whether heterogeneity in CHR can be organized into biologically interpretable brain-symptom dimensions. We applied PLS to identify latent dimensions linking RSFC with diverse clinical symptom profiles and then examined the molecular signatures associated with the corresponding brain patterns (see Figure 1 for study overview). We next tested their associations with independent psychosocial and cognitive measures to assess broader external relevance, and evaluated whether CHR could be clustered into relatively homogeneous biotypes based on these dimensions. Finally, using available Month 2 follow-up data, we examined the short-term longitudinal consistency of the identified brain-symptom dimensions and the clinical and connectivity profiles of the derived biotypes. By integrating clinical, neural, molecular, and longitudinal information, this study aimed to provide a multilevel framework for understanding CHR heterogeneity.

**Figure 1.**
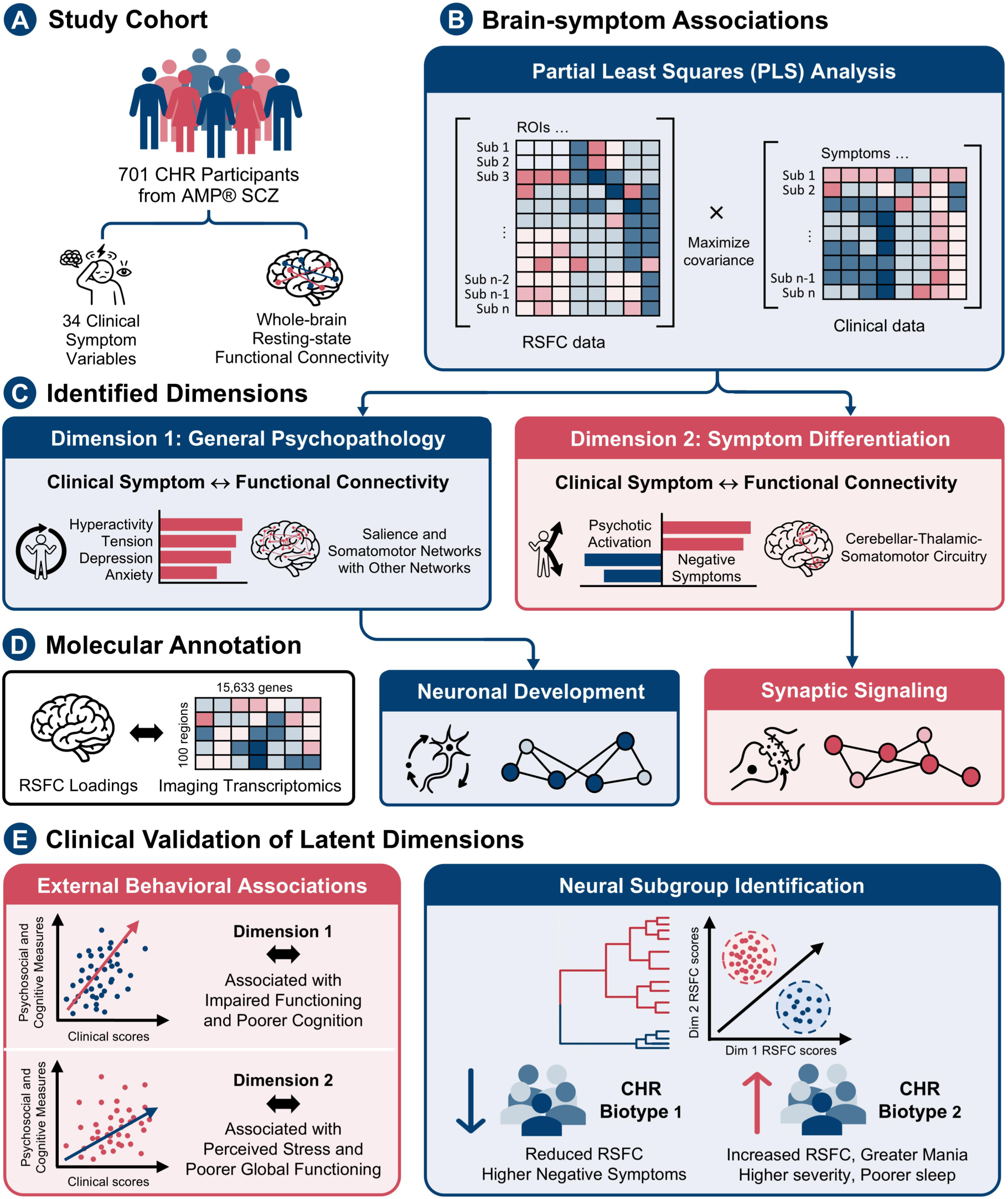
Study overview. (A) Study cohort. Data were drawn from 701 CHR participants in AMP® SCZ, including whole-brain RSFC and 34 clinical symptom measures. (B) Brain-symptom association using PLS. PLS analysis was used to identify latent dimensions that maximize covariance between RSFC and clinical data. (C) Identified brain-symptom dimensions. Two robust latent dimensions were identified, one linking general psychopathology to connectivity between salience and somatomotor networks and other large-scale networks, and the other linking negative symptoms-psychotic activation to cerebello-thalamo-cortical circuitry. (D) Molecular annotation. Transcriptomic decoding showed the general dimension was mainly enriched for neuronal development genes, whereas the symptom-differentiation dimension was mainly enriched for synaptic signaling genes. (E) Clinical validation of latent dimensions. External validation using independent psychosocial and cognitive measures extended the behavioral associations of the latent dimensions. Hierarchical clustering of symptom-related RSFC scores identified two CHR biotypes with distinct RSFC patterns and clinical symptom profiles, supporting the utility of neural subgrouping.

## Methods and Materials

### Participants

We assessed data generated by the Accelerating Medicines Partnership® Schizophrenia Program (AMP® SCZ) (34) and downloaded from the NIMH Data Archive (data release 3.0; doi: 10.15154/81xe-k706). The mission of the AMP Schizophrenia (SCZ) program is to enhance patient treatments by identifying early indicators of schizophrenia. The database (34) includes multimodal imaging and behavioral data from 2,192 participants, including individuals at clinical high risk for psychosis (CHR) and healthy controls (HC), recruited across 42 study sites worldwide. Not all participants have both imaging and behavioral data. All sites adhered to the same standards for assessment and data collection. Participants were aged between 12 and 30 years. CHR participants fulfilled CHR criteria based on the Structured Interview for Prodromal Syndromes (SIPS) and/or the Comprehensive Assessment of at Risk Mental States (CAARMS), whereas healthy controls had no history of serious mental health concerns. Exclusion criteria included intellectual disability, certain nervous system disorders, traumatic brain injury, current or past psychotic disorders, and prior antipsychotic exposure. Details on participant recruitment can be found in the previous AMP Schizophrenia (SCZ) program introduction article (34).

The AMP® SCZ database comprises MRI assessments at baseline and Month 2. Baseline data were used for the primary analyses, with available Month 2 data used for subsequent longitudinal analyses. Baseline MRI data were available for 875 CHR participants, and 838 remained after preprocessing quality control excluding any run that failed quality criteria. After further exclusion of participants with missing data on any clinical or demographic variable, 701 CHR participants were included in the final analyses. Healthy controls were additionally included as a reference sample, with RSFC data available for 244 participants and complete clinical symptom data available for 212 of these participants. Demographic and clinical information are summarized in Supplementary Table S1 and details of missing data are provided in Supplementary Table S2. The study was approved by the governing institutional review board at each participating site and was registered at ClinicalTrials.gov (NCT05905003). All participants provided oral and written informed consent, and the present analyses were conducted using deidentified data.

### Clinical and Behavioral Assessment

The behavioral assessment in AMP® SCZ included a comprehensive range of clinical, cognitive, and psychological measures relevant to the CHR state. A total of 701 participants with complete (non-missing) behavioral data were included in the PLS analysis. The final analysis included 34 symptom-related behavioral variables derived from seven clinical assessment scales: negative symptoms (subscales from the Brief Psychiatric Rating Scale and Negative Symptom Inventory-Psychosis Risk), positive symptoms, mania, and disorientation (subscales from the Brief Psychiatric Rating Scale), depression (subscales from the Brief Psychiatric Rating Scale and the Calgary Depression Scale for Schizophrenia), anxiety (Overall Anxiety Severity and Impairment Scale), sleep quality (8-item PROMIS for Sleep), and global illness severity (Patient Global Impression Scale-Severity). Positive symptom severity assessed with the Positive Symptoms CAARMS Harmonized with SIPS was unavailable for 304 of the 701 participants (43.37%) and was therefore excluded from the primary PLS analysis. Given its clinical relevance, it was further evaluated in control analyses and examined separately in subsequent correlations with the PLS clinical composite scores. See Supplementary Table S1 for details of the measures.

### MRI Acquisition and Preprocessing

Structural and functional MRI data were collected across multiple sites using harmonized MRI protocols implemented on 3T scanners. See Supplementary Methods 1 for details. T1-weighted (T1w) images and resting-state fMRI time series were processed using fMRIPrep (version 24.1.1) (35). T1w images underwent intensity non-uniformity correction with the N4BiasFieldCorrection (ANTs, version 2.3.3), skull stripping via Nipype (version 1.6.1) using the ANTs brain extraction workflow, and tissue segmentation with FSL’s FAST tool (version 5.0.9). Cortical surface reconstruction was performed using FreeSurfer (version 6.0.1). Additionally, the T1w image was non-linearly registered to the MNI152 T1 template using the antsRegistration tool for volume-based spatial normalization.

Functional images were preprocessed using standard pipelines. For functional preprocessing, a skull-stripped reference BOLD volume was created and co-registered with a B0 field map. The susceptibility distortion field was estimated using a pair of spin-echo EPI scans with opposite phase-encoding polarities (AP and PA), as specified in the AMP® SCZ protocol. The corrected BOLD reference was subsequently rigidly co-registered to the T1w reference using FreeSurfer’s bbregister (six degrees of freedom, boundary-based registration). Head motion parameters were estimated using FSL’s mcflirt, generating six rotation and translation parameters. The BOLD time series were further resampled into standard space, aligned with the MNI152NLin6Asym T1 template, using antsApplyTransforms with Lanczos interpolation to minimize smoothing effects. A 6-mm full-width half-maximum Gaussian filter was applied for spatial smoothing. Nuisance regression was performed including the Friston 24-parameter head motion model, as well as mean signals from white matter and cerebrospinal fluid. The data were subsequently temporally band-pass filtered (0.01–0.1 Hz) to retain low-frequency fluctuations relevant to resting-state functional connectivity.

### Resting-State Functional Connectivity

For each participant, resting-state functional connectivity (RSFC) was calculated by concatenating the time series from two runs and computing Pearson’s correlation between the mean time series of 264 regions of interest (200 cortical (36), 32 subcortical (37), and 32 cerebellar (38) regions), resulting in a 264 × 264 connectivity matrix.

### Partial Least Squares

Partial least squares (PLS) is a statistical technique that extracts latent components to relate two sets of multivariate data by maximizing their covariance, thereby achieving an optimal least-squares fit (21,39,40). In our study, PLS was applied to whole-brain RSFC and clinical data from all CHR participants. Age, sex, and education level were regressed out from both RSFC and clinical data, whereas mean framewise displacement (FD) was additionally regressed out from the RSFC data. The resulting data were z-scored across participants, and a cross-covariance matrix between RSFC and clinical variables was computed, followed by singular value decomposition (SVD) to derive latent components (LCs).

Each significant LC corresponds to a unique RSFC pattern (termed RSFC saliences) and a unique clinical pattern (termed clinical saliences). Composite scores for each LC were generated by projecting the RSFC data and clinical variables onto respective saliences of all participants. To estimate the significance of each LC, we performed 10,000 permutation tests, accounting for site effects (sites with fewer than 10 participants were combined), and applied false discovery rate (FDR) correction to LCs explaining more than 5% of the variance (*q* < 0.05).

To further characterize the RSFC and clinical variables associated with each LC, we computed Pearson’s correlations between the original and composite RSFC scores (termed RSFC loadings), as well as between the original and composite clinical scores (termed clinical loadings). A larger absolute value of a loading indicates a greater contribution of the corresponding variable for the LC. To assess the significance of these loadings, we used a bootstrapping procedure with 1000 samples drawn from the original RSFC and clinical data. *P*-values, calculated based on each loading’s position within its respective sample distribution, were all underwent post hoc FDR correction for multiple comparisons (*q* < 0.05).

### Longitudinal Stability

To assess the longitudinal stability of the LCs, we independently repeated the PLS analysis using Month 2 data (*n* = 313) and compared the loading patterns of the corresponding LCs between baseline and Month 2. Pearson correlations were calculated separately for the clinical and RSFC loadings. Loadings were used because they characterize the associations of the original variables with each latent dimension and therefore provide an interpretable measure of pattern correspondence across independently estimated models.

### Control and Reliability Analyses

To assess the robustness of the LCs, we conducted several control analyses (See Supplementary Methods 2 for details). First, we performed a five-fold cross-validation of the PLS analysis. Second, principal component analysis (PCA) was reapplied on the clinical variables. Third, quantile normalization was applied to the clinical variables before PLS analysis. Fourth, the PLS analysis was repeated after excluding a clinical variable with a skewed distribution. Fifth, age, sex, education, and head motion were incorporated as clinical variables and the PLS analysis was repeated. Sixth, ComBat harmonization was applied to the RSFC data to account for potential site effects. Seventh, the PLS analysis was repeated in participants with available Positive Symptoms CAARMS Harmonized with SIPS (PSYCHS) data after adding positive symptom severity to the clinical variables. Eighth, as evidence that the global fMRI signal may carry clinically meaningful information in psychosis and that its removal may attenuate disease related variance (18,41), global signal regression (GSR) was not included in the primary preprocessing pipeline. To address ongoing debate over GSR, we repeated the PLS analysis after GSR and examined associations between global BOLD fluctuation amplitude (42) and the LC1 and LC3 composite scores. In addition to the five-fold cross-validation in the first step, robustness in all subsequent steps was evaluated by correlating brain RSFC and clinical saliences from each control analysis with those from the original PLS model.

### Gene Expression Pattern Associated with RSFC Loadings

To characterize RSFC-symptom associations in relation to molecular features, we derived regional RSFC loadings for each ROI by summing the absolute RSFC loadings with all other ROIs. Whole-brain gene expression profiles were obtained from the Allen Human Brain Atlas (AHBA; https://human.brain-map.org/) (43) transcriptomic dataset, following standardized preprocessing (https://github.com/BMHLab/AHBAprocessing; Supplementary Table S3) (26). See Supplementary Methods 3 for details. Because right-hemisphere coverage in the AHBA is limited, analyses were restricted to the left hemisphere of the Schaefer atlas (26), comprising 100 regions and 15,633 genes.

PLS regression (44) was applied to examine associations between regional RSFC loadings and transcriptional profiles, with gene expression as predictors and RSFC loadings as responses. The first PLS component (PLS1) captured the cortical gene expression pattern most strongly associated with RSFC loadings. The spatial correlation between the PLS1 scores and RSFC loadings was assessed, with spatial autocorrelation accounted for using a spin-based permutation test (5,000 rotations). Gene weights were assessed by bootstrapping and expressed as Z-scores. Genes surviving FDR correction (*q* < 0.01) were divided into PLS1+ (Z > 0) and PLS1− (Z < 0) gene lists for subsequent enrichment analyses.

### Gene Enrichment and Specific Expression Analyses

To investigate the potential molecular impact underlying RSFC loadings, functional enrichment analyses were performed using Metascape (http://metascape.org/)(45) to identify GO biological processes and KEGG pathways. PLS1+ and PLS1− gene lists were analyzed separately, with significance defined at FDR correction (*q* < 0.01).

### Associations of Clinical Dimensions with Psychosocial and Cognitive Measures

As a post hoc assessment of external validity, we examined whether the clinical dimensions identified by PLS extended to psychosocial and cognitive domains. To maintain the primary focus on clinical symptom heterogeneity, these broader measures were examined separately as external correlates. These measures were selected to capture clinically relevant domains, including positive symptom severity (Positive Symptoms CAARMS Harmonized with SIPS), suicidal intensity (Columbia-Suicide Severity Rating Scale), perceived discrimination (Perceived Discrimination Scale), perceived stress (Perceived Stress Scale), global functioning (Social and Occupational Functioning Assessment Scale), role, and social functioning (subscales from the Global Functioning), as well as cognitive performance assessed by the PennCNB (see Supplementary Table S4 for details). Pearson correlations were used to examine associations between the clinical composite score for each significant LC and each external measure. Analyses were conducted using available participants for each measure, resulting in varying sample sizes. Statistical significance was determined using the Benjamini–Hochberg false discovery rate (FDR) correction applied across all correlations (*q* < 0.05).

### Hierarchical Clustering based on RSFC Dimensions

To further explore the implications of symptom-informed brain dimensions, brain imaging data were used to cluster CHR participants into distinct biotypes. Specifically, we focused on RSFC composite scores from the significant PLS latent components (i.e., LC1 and LC3), which showed robust associations in the primary analysis. First, a subject-by-subject dissimilarity matrix was constructed using Euclidean distance in a two-dimensional space defined by these LCs. Then, hierarchical clustering was performed using Ward’s linkage method, which iteratively merges clusters to minimize the increase in within-cluster variance and produces a hierarchical cluster tree (31,46). The optimal number of clusters was determined by inspecting the dendrogram and evaluating the average silhouette index (31), with the Calinski-Harabasz criterion used as an additional measure of clustering performance (47). Clustering stability was assessed by repeating hierarchical clustering in two-thirds subsamples (1,000 replicates) and comparing each subsample clustering with the full-sample clustering using adjusted mutual information (AMI) (47). Stability was summarized as the mean AMI, and its significance was assessed using a null distribution generated from 1,000 permutations. To characterize the clinical and neural features of the identified CHR biotypes, clinical symptoms (see Supplementary Table S1) and whole-brain RSFC were compared between the two biotypes, with healthy controls included as a reference group. For the RSFC comparisons, we examined how the latent biotypes were represented in the original connectivity space, with findings interpreted descriptively rather than as an independent validation of neurobiological distinctiveness. Group differences were assessed using one-way analysis of variance (ANOVA), followed by post hoc pairwise comparisons using Welch’s two-sample *t*-tests to account for unequal sample sizes. For each pairwise comparison, multiple comparisons were corrected separately using the Benjamini–Hochberg FDR (*q* < 0.05).

Additionally, we evaluated the longitudinal consistency of the CHR biotypes using participants with both baseline and Month 2 data, with RSFC and clinical symptoms examined separately. For RSFC, pairwise group differences at Month 2 were assessed using Welch’s two-sample *t*-tests, and their similarity to baseline was evaluated by Pearson correlations between the corresponding *t*-value patterns. Edgewise connection change scores were calculated as Month 2 minus baseline and compared between groups using Welch’s two-sample *t*-tests. The same analyses were applied to the clinical symptoms. Multiple comparisons were corrected separately within each group comparison using the Benjamini–Hochberg FDR procedure (*q* < 0.05).

## Results

### PLS Reveals Two Robust LCs Linking Symptoms and RSFC

PLS analysis was conducted on whole-brain RSFC and 34 clinical variables from 701 participants, with age, sex, education, and head motion included as regressors. LC1 and LC3 were significant after FDR correction, whereas LC2 did not reach significance in permutation testing (*p* = .36) and was not interpreted further. To evaluate longitudinal stability, we repeated the PLS analysis using Month 2 data. The Month 2 loading patterns were strongly correlated with the baseline loading patterns for both LC1 (clinical, *r* = 0.76, *p* < 0.001; RSFC, *r* = 0.87, *p* < 0.001) and LC3 (clinical, *r* = 0.75, *p* < 0.001; RSFC, *r* = 0.72, *p* < 0.001), indicating longitudinal stability of both dimensions.

Five-fold cross-validation yielded significant correlations between clinical and RSFC composite scores in the test folds for LC1 (mean *r* = 0.12, all *p* < 0.001) and LC3 (mean *r* = 0.20, all *p* < 0.001), supporting the generalizability of the identified brain-symptom associations. Control and reliability analyses generally showed moderate to high correspondence with the primary PLS patterns after quantile normalization, exclusion of the skewed clinical variable, covariate inclusion, ComBat harmonization, inclusion of PSYCHS positive symptom severity, and GSR (all *p* < 0.01; see Supplementary Table S5 for details). For GSR, correspondence with the primary PLS patterns varied across dimensions, while the association between global BOLD fluctuation amplitude and the RSFC composite score was higher for LC1 than LC3, potentially suggesting greater sensitivity of LC1 to global signal variation (see Supplementary Results for details).

### LC1 Reflects General Psychopathology

The first latent component (LC1) explained 24% of the covariance between RSFC and clinical symptoms, with a significant correlation (*r* = 0.20, *p* = 0.001) between RSFC and clinical composite scores (Figure 2A). To interpret the contributions of RSFC and clinical symptoms to LC1, we computed Pearson’s correlations between the original and composite clinical scores (i.e., clinical loadings), with larger absolute values indicating greater contributions. Higher clinical composite scores were associated with more severe general psychopathology (e.g., motor hyperactivity, tension, and depression) and poorer sleep quality. The top 20 correlations are shown in Figure 2B.

**Figure 2.**
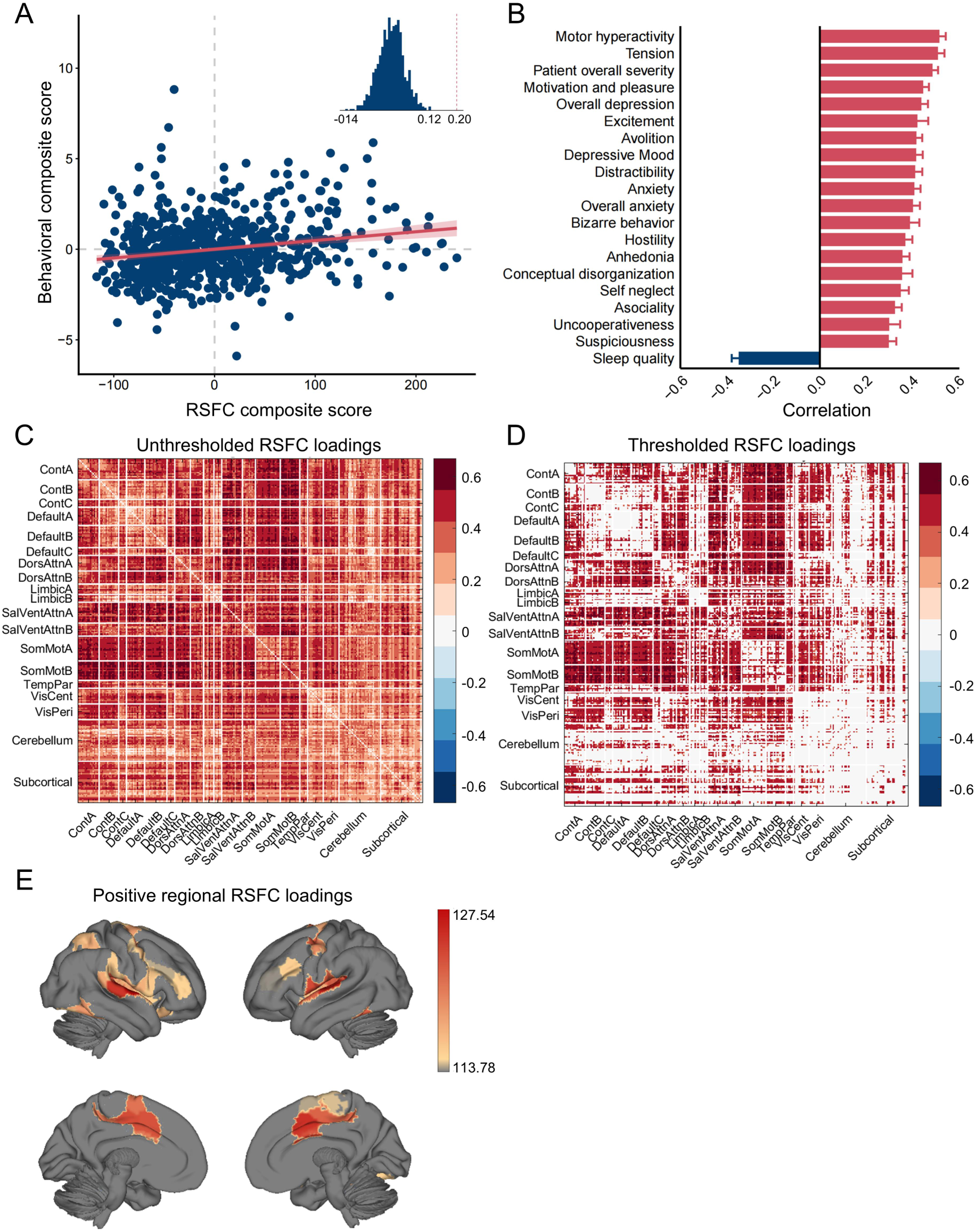
First latent component (LC1) reflects general psychopathology. (A) Correlation between RSFC and clinical composite scores in CHR participants (*r* = 0.20, *p* = 0.001). The inset presents the null distribution obtained from permutation testing, with the dashed line marking the observed correlation. (B) Top 20 correlations between the original and composite clinical scores of LC1 (i.e., clinical loadings). Error bars indicate bootstrapped standard deviation. (C) Unthresholded correlations between the original and composite RSFC metrics (i.e., RSFC loadings). (D) Thresholded correlations between the original and composite RSFC metrics (FDR, *q* < 0.05). Only connections with absolute values greater than 0.4 are displayed to maintain visualization clarity. (E) Regional positive RSFC loadings, computed by summing positive RSFC loadings of each ROI with all other regions, for the top 20% of regions.

Figure 2C (unthresholded correlations) and Figure 2D (significant correlations) present the correlations of LC1 between original and composite RSFC metrics across 200 cortical, 32 cerebellar, and 32 subcortical regions (i.e., RSFC loadings). Higher RSFC composite scores were associated with increased RSFC of the somatomotor and salience A networks with the control, default mode, and dorsal attention A networks. Regional RSFC loadings were computed for each ROI by summing its positive loadings with all other regions, given that RSFC patterns were predominantly positive (Figure 2E). For LC1, regional loadings were predominantly positive across the cortex, with the strongest effects (top 20% of regional loadings) mainly concentrated within the salience network (e.g., middle cingulate cortex, rolandic operculum and insula) and somatomotor network (e.g., precentral and postcentral gyri).

### LC3 Reflects Negative Symptoms-Psychotic Activation Dimension

The third latent component (LC3) explained 10% of the covariance between RSFC and clinical symptoms, with a significant correlation (*r* = 0.33, *p* = 0.001) between RSFC and clinical composite scores (Figure 3A). Higher clinical composite scores were positively associated with more severe negative symptoms (e.g., anxiety and depression) and negatively associated with elevated psychotic activation (e.g., conceptual disorganization and bizarre behavior; Figure 3B). Correlations between original and composite RSFC metrics were increased for the cerebellar and subcortical regions with other networks, and decreased within the somatomotor network (Figures 3C and 3D). Regional RSFC loadings for LC3 showed positive values mainly in subcortical and cerebellar regions (e.g., thalamus and cerebellar vermis), while negative values were primarily observed in the somatomotor network (e.g., postcentral gyrus; Figures 3E and 3F).

**Figure 3.**
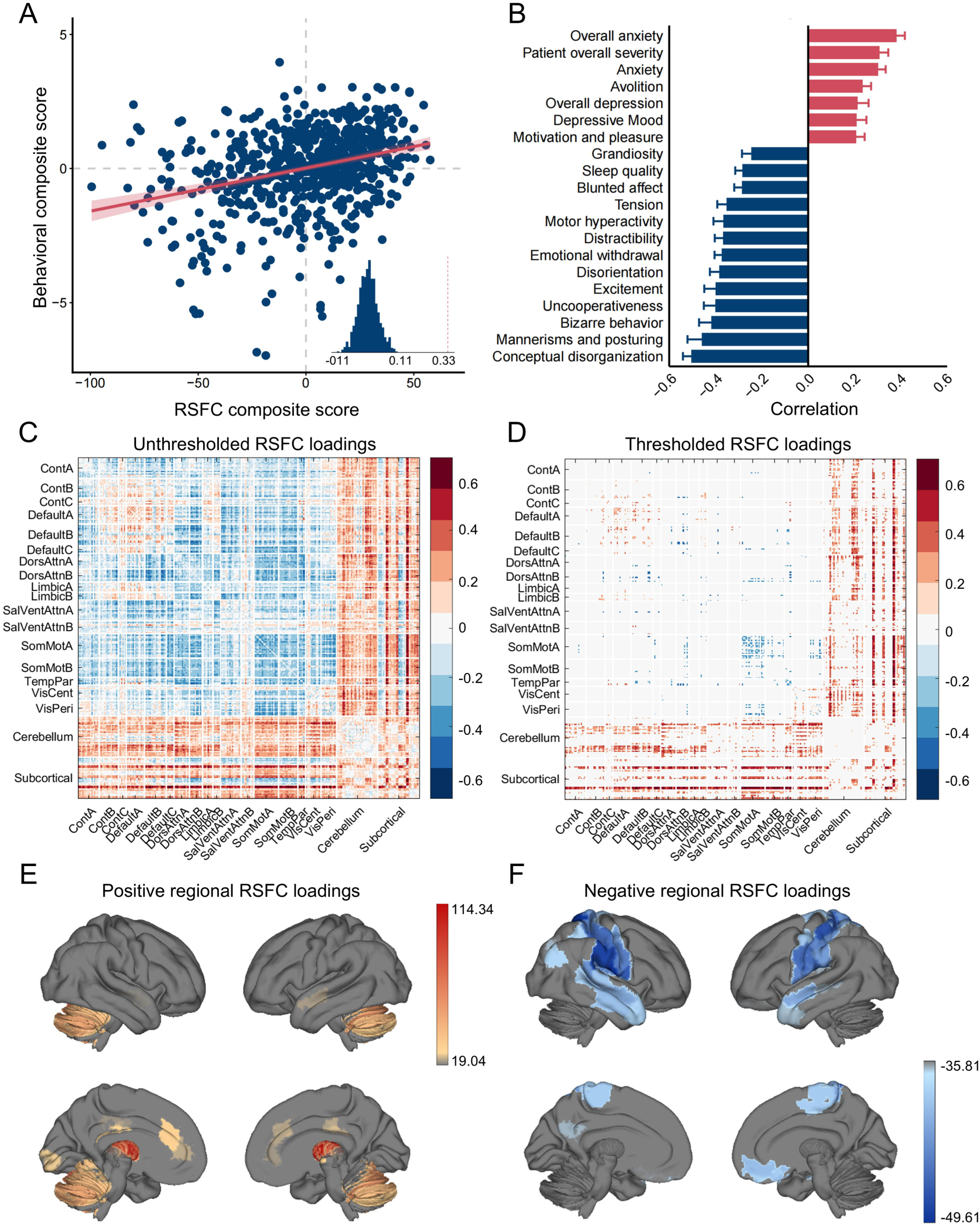
Third latent component (LC3) reflects negative symptoms-psychotic activation dimension. (A) Correlation between RSFC and clinical composite scores in CHR participants (*r* = 0.33, *p* = .001). The inset presents the null distribution obtained from permutation testing, with the dashed line marking the observed correlation. (B) Top 20 correlations between the original and composite clinical scores of LC3 (i.e., clinical loadings). Error bars indicate bootstrapped standard deviation. (C) Unthresholded correlations between the original and composite RSFC metrics (i.e., RSFC loadings). (D) Thresholded correlations between the original and composite RSFC metrics (FDR, *q* < 0.05). (E) Regional positive RSFC loadings, computed by summing positive RSFC loadings of each ROI with all other regions, for the top 20% of regions. (F) Regional negative RSFC loadings, computed by summing negative RSFC loadings of each ROI with all other regions, for the top 20% of regions.

### Functional and Expression Characterization of RSFC Loadings in LC1 and LC3

PLS regression was used to identify transcriptomic patterns associated with regional RSFC loadings, computed as the sum of absolute RSFC loadings with all other ROIs. The first PLS component (PLS1; Figures 4A and Supplementary Figure S2) revealed a gene expression pattern that is strongly associated with regional RSFC loadings, explaining 36.53% of the variance for LC1 (*p* = 0.001) and 22.07% for LC3 (*p* = 0.03).

**Figure 4.**
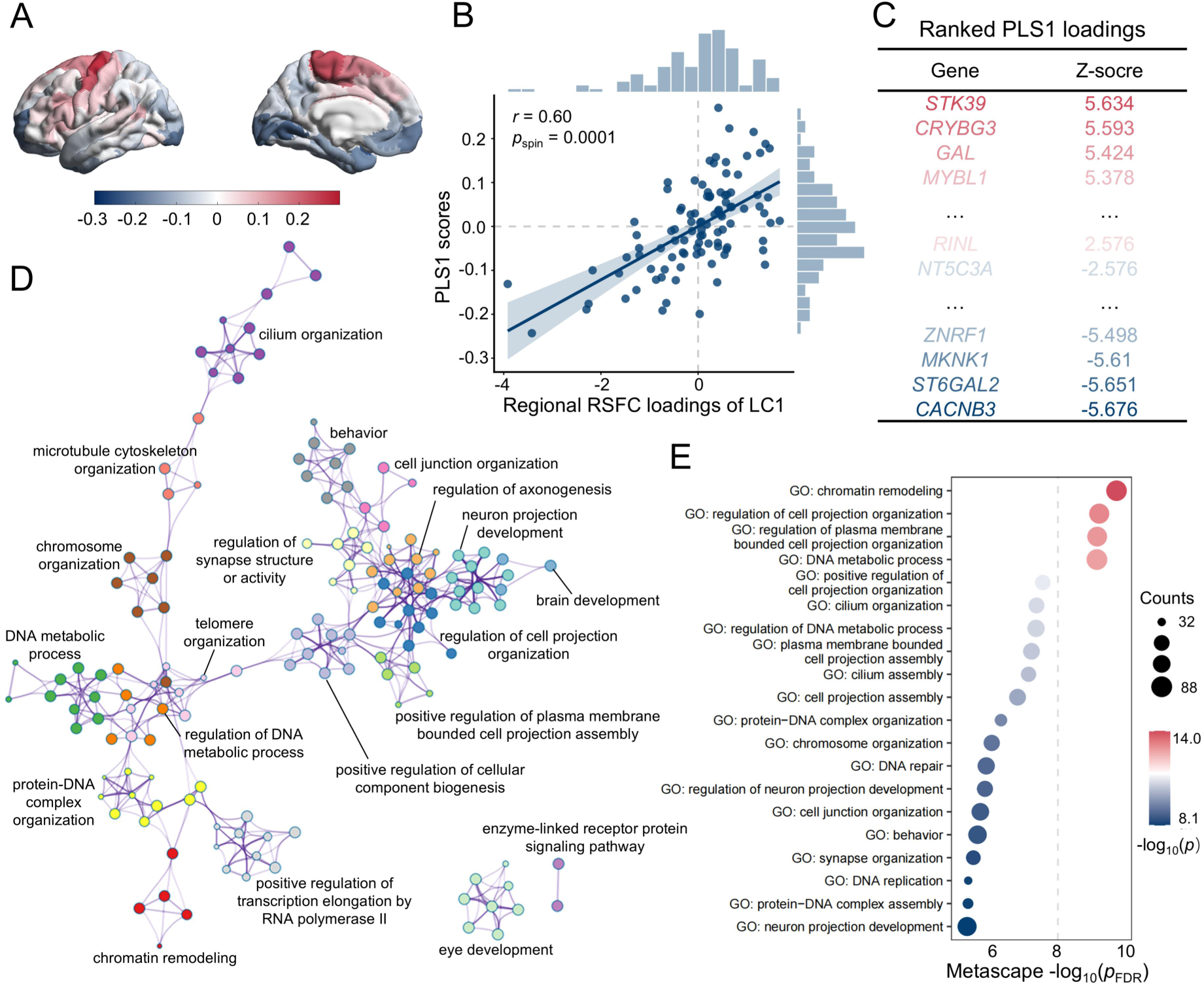
Transcriptomic decoding of regional RSFC loadings in LC1. (A) Cortical distribution of regional PLS1 scores in the left hemisphere. (B) Association between RSFC loadings of LC1 and PLS1 scores (*r* = 0.60, spin-test *p* = 0.0001). (C) Ranked PLS1 loading. (D) The Metascape enrichment network shows intra- and inter-cluster similarities among enriched terms. Node size reflects the number of input genes, and node color indicates cluster membership. (E) Enrichment analysis of genes transcriptionally associated with RSFC loadings of LC1 (FDR, *q* < 0.01). Circle size indicates the number of genes associated with each pathway.

For LC1, a significant spatial association was observed between RSFC loadings and PLS1 scores (*r* = 0.60, *p*spin = 0.0001; Figure 4B). Meanwhile, functional annotation of PLS-derived genes was performed using Metascape, including enrichment analyses of GO biological processes and KEGG pathways. After FDR correction (*p* < 0.01), 1,894 positively weighted (PLS1+) and 1,478 negatively weighted (PLS1−) genes were identified. Enrichment analysis showed that PLS1− genes were mainly enriched in biological processes related to neuronal development, synaptogenesis, DNA replication, and DNA repair (Figures 4C−4E), whereas PLS1+ genes were primarily enriched in ion transport and nervous system development (Supplementary Figure S1).

For LC3, RSFC loadings showed a significant spatial correlation with PLS1 scores (*r* = 0.47, spin-test *p* = 0.0002). FDR correction identified 1,386 positively and 1,222 negatively weighted genes. PLS1− genes were mainly enriched in synaptic signaling and neuron projection development (Supplementary Figures S2), whereas PLS1+ genes were primarily associated with RNA processing, protein modification, and cell cycle processes (Supplementary Figure S3).

### Associations Between Clinical Composite Scores and Psychosocial and Cognitive Measures

Following the identification of two RSFC-symptom dimensions, we evaluated their psychosocial and cognitive relevance by examining associations between clinical composite scores and standardized independent measures. LC1, reflecting general psychopathology, showed significant associations with positive symptom severity (*r* = 0.36, *p*FDR < 0.001), suicidal intensity (*r* = 0.17, *p*FDR < 0.001), perceived discrimination (*r* = 0.18, *p*FDR < 0.001), perceived stress (*r* = 0.44, *p*FDR < 0.001), global functioning (*r* = −0.31, *p*FDR < 0.001), role functioning (*r* = −0.21, *p*FDR < 0.001) social functioning (*r* = −0.23, *p*FDR < 0.001) and cognitive performance in the PennCNB Digit Symbol test (*r* = −0.15, *p*FDR = 0.02; Figure 5A). LC3, representing a negative symptom-psychotic activation dimension, was significantly associated with perceived stress (*r* = 0.26, *p*FDR < 0.001) and global functioning (*r* = −0.14, *p*FDR = 0.0021; Figure 5B).

**Figure 5.**
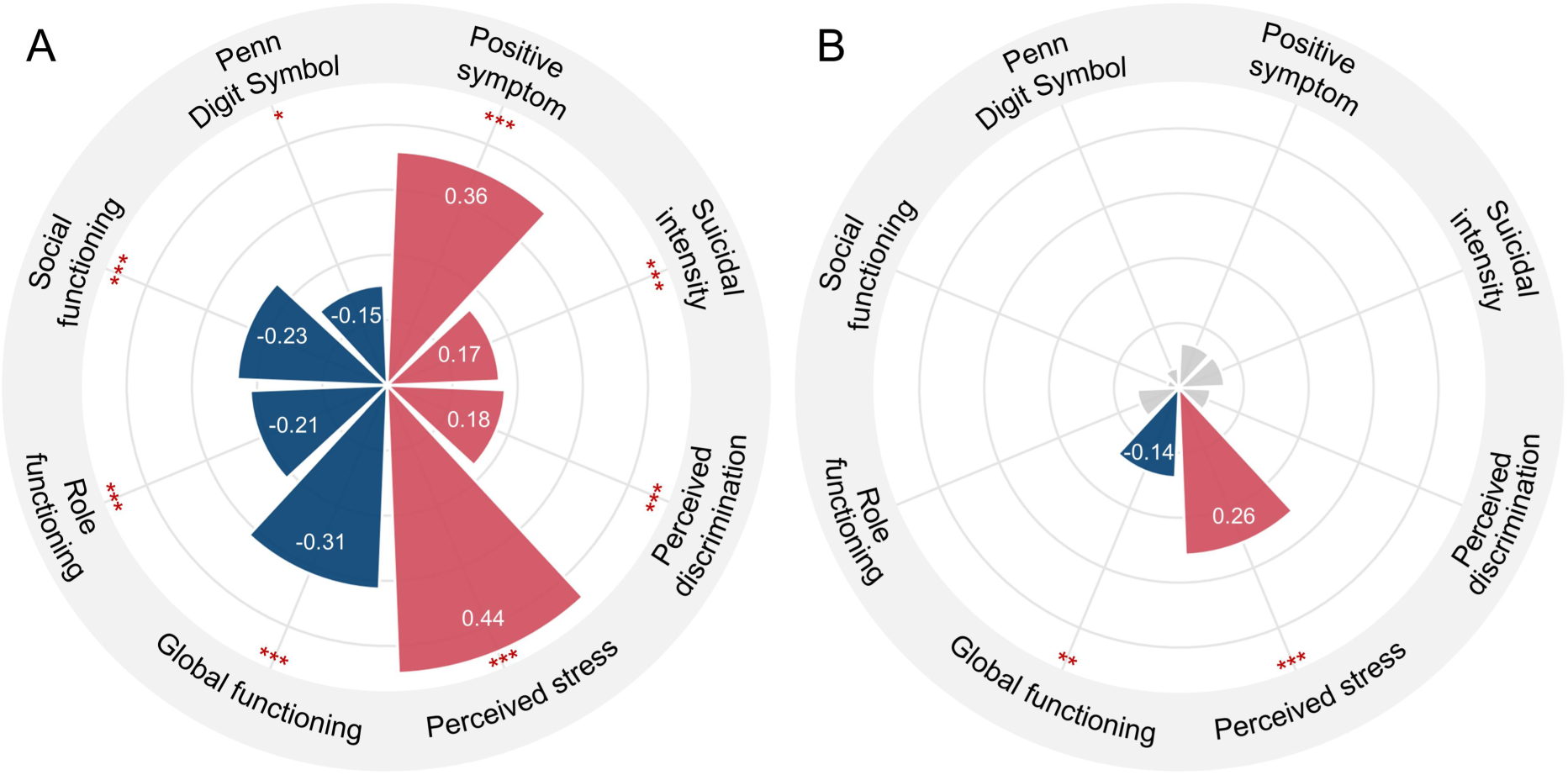
Associations between clinical composite scores and psychosocial measures. (A) LC1 (general psychopathology) associated positively with positive symptom, suicidal intensity, perceived discrimination, and stress, and negatively with global, role, and social functioning, as well as PennCNB Digit Symbol performance (number of correct responses). (B) LC3 (negative symptom-psychotic activation) associated positively with stress and negatively with global functioning. Deep pink represents significant positive correlations, deep blue represents significant negative correlations, and grey represents non-significant correlations. * *p*FDR < 0.05, ** *p*FDR < 0.01, *** *p*FDR < 0.001.

### Two CHR Biotypes Identified Based on RSFC-symptom Dimensions

To further examine the implications of the two RSFC symptom dimensions in brain space, RSFC composite scores from LC1 and LC3 were used for hierarchical clustering to identify relatively homogeneous biotypes among CHR participants. The number of clusters was evaluated by comparing the average silhouette index and Calinski-Harabasz criterion across 2 to 10 clusters. The silhouette index was highest for two clusters, whereas the Calinski-Harabasz values were comparable for two and three clusters. Considering both criteria, two clusters were retained (Supplementary Figure S4A and B), comprising 577 individuals in biotype 1 and 124 individuals in biotype 2 (Figure 6A). Clustering was stable across subsamples, with concordance significantly greater than expected by chance (*p* = 0.001; Supplementary Figure S4).

**Figure 6.**
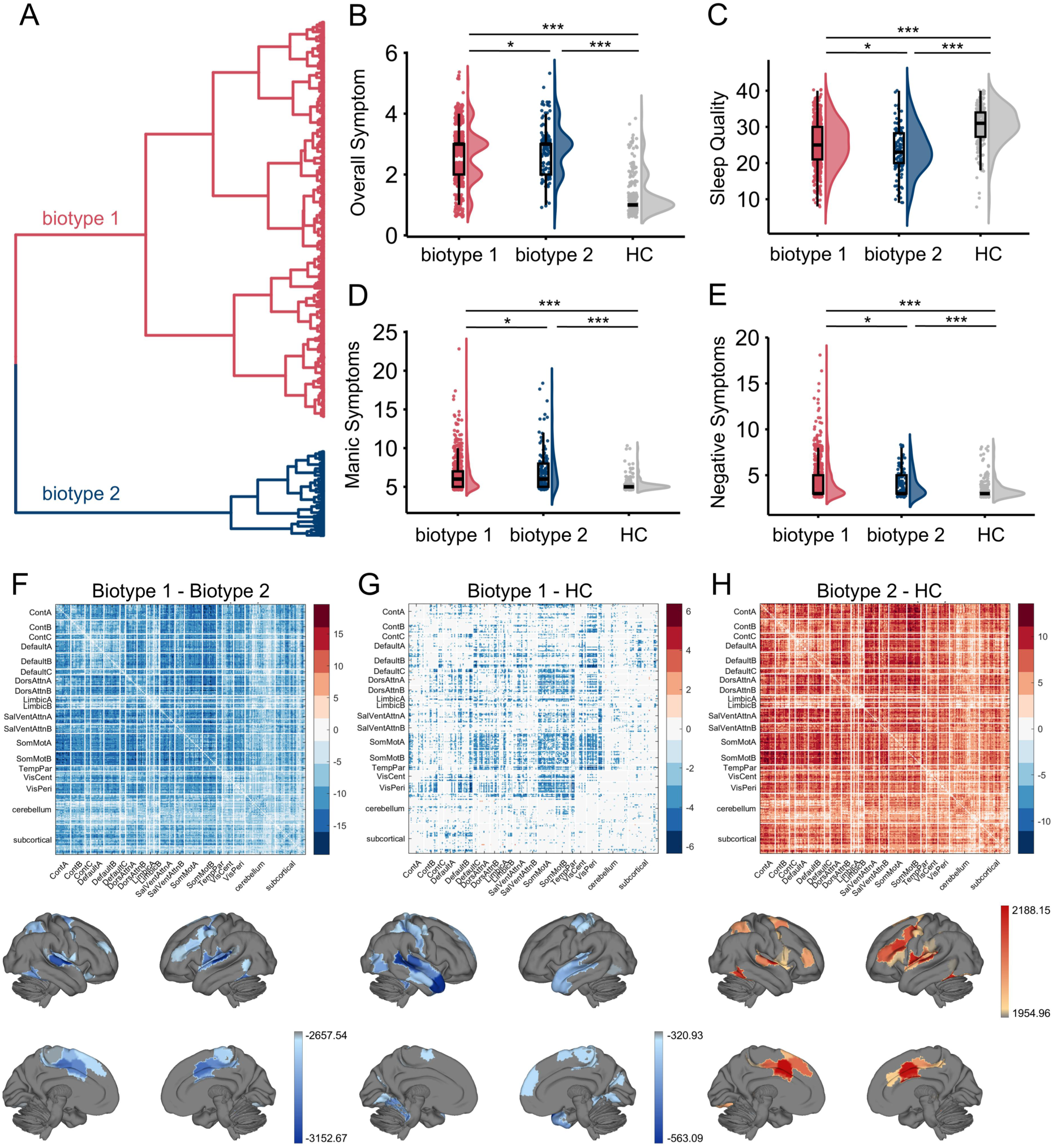
Clinical symptom and Whole-brain RSFC differences across two CHR biotypes and healthy controls. (A) Hierarchical clustering of RSFC composite scores using Ward’s linkage identified two CHR biotypes (*n*biotype 1 = 577, *n*biotype 2 = 124). Dendrogram linkage heights represent the distance between clusters. (B–E) Clinical symptoms comparisons across the two biotypes and healthy controls (HC). Biotype 1 exhibited higher negative symptoms, whereas biotype 2 showed higher manic symptoms, greater overall symptom severity, and poorer sleep quality. HC showed lower symptom severity, negative symptoms, and manic symptoms, and better sleep quality compared with both biotypes. * *p*fdr < 0.05, ** *p*fdr < 0.01, *** *p*fdr < 0.001. (F-G) Whole-brain RSFC comparisons across the two biotypes and HC. Biotype 1 exhibited predominantly reduced RSFC relative to both Biotype 2 and HC, whereas Biotype 2 showed predominantly increased RSFC compared with HC. Top panels display edge-wise RSFC *t*-value matrices (FDR, *q* < 0.05), and bottom panels show regional summaries, computed for each ROI by summing the positive or negative RSFC *t*-values across all its connections (FDR, *q* < 0.05), for the top 20% of regions.

To characterize the clinical profiles of the two biotypes and their RSFC patterns in the original RSFC space, we compared Biotype 1 and Biotype 2 and assessed each biotype relative to healthy controls. For behavioral analyses, significant group differences were observed across several clinical symptoms (Figures 6B−6E). Biotype 1 exhibited higher negative symptom scores than biotype 2 (*t* = 2.38, *p*FDR = 0.045), whereas biotype 2 showed higher manic symptoms (*t* = −2.74, *p*FDR = 0.035), greater self-reported overall symptom severity (*t* = −2.73, *p*FDR = 0.035), and poorer sleep quality (*t* = 2.40, *p*FDR = 0.045). Compared with both biotypes, HC showed significantly lower overall symptom severity, negative symptoms, and manic symptoms, and better sleep quality.

Whole-brain RSFC also differed significantly among the three groups (Biotype 1, Biotype 2, and HC; Figures 6F−6H). Compared with Biotype 2, Biotype 1 exhibited predominantly reduced RSFC, with decreases primarily observed in SomMot-Cont, SomMot-attention, SomMot-Default, and Default-attention connections. At the regional level, negative RSFC *t*-values, calculated by summing negative connectivity across all connections for each ROI, were most pronounced (top 20%) in the somatomotor network (e.g., superior temporal gyrus and Heschl’s gyrus). Relative to healthy controls, Biotype 1 showed decreased RSFC, whereas Biotype 2 exhibited increased RSFC.

### Longitudinal Consistency of Biotype Differences

To further evaluate the longitudinal stability of the biotypes identified at baseline, clinical and RSFC differences were examined at Month 2. Clinical differences between the two biotypes were highly consistent between baseline and Month 2 (*r* = 0.85, *p* = 0.002), with similar consistency observed for comparisons of each biotype with HC (Supplementary Figure S5A–S5C). Changes from baseline to Month 2 in sleep quality differed significantly between the two biotypes before correction (*t* = −2.67, *p* = .009), but did not survive FDR correction (*p*FDR = .085; Supplementary Figure S5D). Relative to HC, Biotype 1 showed significant differences in changes in depressive symptoms, anxiety, and positive symptoms, whereas Biotype 2 differed in changes in sleep quality and positive symptoms (Supplementary Figure S5E–S5F).

RSFC differences were also consistent across time. The *t*-value patterns distinguishing Biotype 1 from Biotype 2 were positively correlated between baseline and Month 2 (*r* = 0.66, *p* < 0.001), with similar consistency observed in comparisons with HC (Supplementary Figure S6). At Month 2, Biotype 1 continued to show predominantly lower RSFC than Biotype 2 (Supplementary Figure S7). Biotype 2 also showed predominantly higher RSFC than HC, whereas no connections between Biotype 1 and HC survived FDR correction. Longitudinal change analyses further showed greater increases in RSFC in Biotype 1 than in Biotype 2 and greater decreases in Biotype 2 than in HC, whereas no differences in RSFC change between Biotype 1 and HC survived FDR correction (Supplementary Figure S8).

## Discussion

The present study suggests that heterogeneity in CHR is not merely diffuse variation across symptoms and neural features, but can be organized along separable brain-symptom dimensions with distinct levels of clinical specificity and biological annotation. In a large multisite CHR cohort, we identified one dimension indexing broad psychopathology-related dysfunction and another capturing symptom differentiation within the psychosis risk state. These dimensions were reproducible at Month 2 and were further supported by transcriptomic annotation, external psychosocial and cognitive correlates, and longitudinally consistent dimension-informed biotypes, together providing a multilevel account of CHR heterogeneity.

LC1 appeared to index a general psychopathology burden dimension, capturing a broad source of clinical heterogeneity in CHR. Its convergence with the transdiagnostic *p*-factor suggests that CHR encompasses general psychopathology beyond psychosis risk symptoms (48,49). Building on prior evidence from symptom structure, our findings further identify this pattern through the covariation between clinical features and brain connectivity (49). At the neural level, the most prominent alterations were observed in functional connectivity between the salience and somatomotor networks and other large-scale networks, including control, default mode, and attentional systems. The salience network plays a central role in detecting and integrating internal and external information and in coordinating switching between internally and externally oriented brain states (50–52). Its dysfunction has been implicated across the psychosis spectrum, including in CHR individuals (15), first-episode psychosis (53), and chronic schizophrenia (51), suggesting that salience network abnormalities may emerge early and represent a potential vulnerability marker for psychosis. The somatomotor network, although traditionally associated with sensorimotor processing, has also been increasingly linked to broader psychopathology dysfunction (17). In psychosis, functional dysconnectivity of somatomotor networks is associated with the severity of neural impairment (54) and has been proposed as a robust biomarker (55). Taken together, these findings suggest that general psychopathology in CHR may be preferentially associated with dysregulated interactions between higher-order networks and salience and somatomotor systems, rather than with isolated dysfunction within a single network. This interpretation is further supported by the broader clinical correlates of LC1, which extended beyond symptom burden and sleep disturbance to impaired functioning and worse cognitive performance. The greater sensitivity of LC1 to global signal treatment, together with its stronger association with global BOLD fluctuation amplitude, suggests that the broad psychopathology related covariance captured by LC1 partly overlaps with individual differences in global BOLD fluctuations, whereas LC3 showed comparatively less dependence on global signal variation. Because the global signal contains both neural and nonneural sources, the basis of this overlap remains to be clarified. Nevertheless, these findings highlight the potential clinical relevance of global signal variation (18,41) and motivate further investigation in relation to specific symptom dimensions.

In contrast to LC1, LC3 appeared to capture a more specific dimension of symptom configuration within the CHR state. At the behavioral level, this dimension spanned greater motivational and internalizing features at one pole and more pronounced overactivation and disorganization at the other, suggesting variation in how psychopathology is expressed across individuals at risk for psychosis (7,56). At the neural level, LC3 was characterized primarily by altered connectivity involving cerebellar and subcortical regions, particularly the thalamus and cerebellar vermis, together with reduced connectivity within the somatomotor network. This pattern aligns with established evidence implicating cerebello-thalamo-cortical (CTC) dysconnectivity across the psychosis spectrum (57–59). Previous studies have consistently demonstrated that CTC hyperconnectivity is robustly observed across diverse fMRI paradigms in both clinical and high-risk populations, suggesting its potential as a state-independent neural signature for psychosis prediction and characterization (57,58). Mechanistically, such hyperconnectivity may represent a downstream phenomenon of cortical inhibitory deficits (58,60,61). Alternatively, it may reflect a compensatory effort by the cerebellum to counteract excessive error input from the upstream cerebral cortex in order to coordinate disorganized thoughts and behaviors (58,62). Compared with LC1, LC3 therefore appeared less reflective of broad nonspecific burden and more indicative of how symptom profiles are configured within the psychosis risk state. This interpretation is supported by its more selective external associations, as LC3 was primarily related to perceived stress and poorer global functioning. Taken together, these findings suggest that LC3 captures a more circumscribed form of clinical heterogeneity in CHR, linking symptom differentiation to CTC dysconnectivity.

The transcriptomic findings place the present dimensions in the context of biological themes that have been repeatedly implicated in psychosis research (63,64). Prior work has consistently implicated neurodevelopmental disturbance and synaptic dysfunction as two major biological processes relevant to psychosis vulnerability (64–66). Against this background, the present findings are notable in suggesting that these processes may show differential correspondence with dimensions of heterogeneity within CHR, rather than being uniformly represented across the risk state. Specifically, LC1-related cortical patterns showed enrichment for genes related to neuronal development, synaptogenesis, and DNA replication and repair, broadly consistent with prior evidence implicating developmental vulnerability (64,67) and impaired cellular maintenance (68,69) in schizophrenia-spectrum conditions. By contrast, LC3-related patterns showed enrichment for genes involved in synaptic signaling and neuron projection development, aligning more closely with prior models emphasizing altered synaptic organization and circuit-level information processing in psychosis (65,70). Thus, rather than identifying a single molecular substrate for CHR, our findings suggest that the dysconnectivity patterns associated with broad psychopathology and symptom differentiation show partially distinct transcriptomic profiles. Notably, the transcriptomic findings should be interpreted as molecular annotation rather than evidence of underlying molecular mechanisms (63). Even so, they extend previous work by suggesting that developmental and cellular maintenance processes, as well as synaptic and projection processes, are differentially represented across separable brain-symptom dimensions within the psychosis risk state.

The biotype findings provide a complementary perspective on CHR heterogeneity by organizing the multivariate variation captured jointly by LC1 and LC3 into distinct subgroups. This is notable as prior CHR research has often characterized symptomatic (12,71) or neural (22) profiles independently, with these perspectives traditionally operating in parallel. Our findings extend this literature by highlighting the potential value of deriving subgroups from latent brain-symptom dimensions, which may offer a more integrated insight beyond clinical or connectivity measures alone. Within this framework, Biotype 1 was characterized by reduced RSFC and more prominent negative symptoms, whereas Biotype 2 showed increased RSFC together with greater manic symptoms, higher overall symptom severity, and poorer sleep quality. Relative to healthy controls, the two biotypes also showed divergent connectivity profiles, with functional reduction predominating in Biotype 1 and hyperconnectivity in Biotype 2. Specifically, the most pronounced biotype differences were observed in connectivity between the somatomotor network and other higher-order systems. The observed clinical differentiation suggests that variation along these latent dimensions can give rise to clinically interpretable subgroups, with negative symptoms and more generalized symptom burden characterizing the two biotypes and broadly overlapping with previous CHR clustering findings (71,72). This clinical distinction may therefore extend to the neural level, with evidence linking somatomotor dysconnectivity to clinical heterogeneity in psychosis (22,54,55) suggesting that somatomotor connectivity may help distinguish the two biotypes.

The longitudinal analyses provided additional support for the stability of the identified biotypes, as both clinical and RSFC group difference patterns remained similar at Month 2. This is consistent with previous CHR studies showing longitudinal consistency in distinct symptom and functional trajectories over time (73,74). At the same time, longitudinal clinical course may vary according to symptom profile (7). Negative symptom predominant presentations show greater symptom stability and lower conversion risk, whereas more severe attenuated positive symptoms are associated with higher conversion risk and more rapid onset of psychosis (75). Notably, the present findings further suggest that relatively stable clinical differentiation may coexist with divergent short-term RSFC changes in CHR. However, with follow-up limited to Month 2, it remains unclear whether these early clinical and connectivity differences are associated with longer-term outcomes, including transition to psychosis, remission, or functional recovery. Taken together, dimensional analyses capture shared variation across individuals, whereas categorical clustering organizes this variation into clinically interpretable subgroups.

Several limitations should be noted for the present study. First, the transcriptomic analyses were based on normative spatial gene-expression maps and should therefore be interpreted as molecular annotation rather than direct evidence of CHR specific mechanisms. Second, due to significant missing data on medication use, the potential effects of medication could not be assessed. Third, the present study leveraged the Accelerating Medicines Partnership® Schizophrenia Program (AMP® SCZ(34)) program, the largest multicenter CHR cohort to date, with harmonized protocols for data acquisition and processing across sites. This design strengthens confidence in the observed patterns, although independent replication will still be important for establishing the robustness and broader generalizability of the identified dimensions and biotypes. Fourth, baseline connectivity differences between biotypes partly reflect the RSFC dimensions used to derive the clusters and should therefore be interpreted in this context rather than as independent validation. The similar clinical and RSFC patterns observed at Month 2 provide longitudinal support for their reproducibility, although replication in independent cohorts remains necessary.

In summary, the present findings suggest CHR heterogeneity can be organized into separable and reproducible brain-symptom dimensions that capture both broad psychopathology and more specific variation in symptom configuration. These dimensions link distributed functional connectivity with distinct clinical profiles and transcriptomic signatures, and further delineate clinically interpretable biotypes that show short-term longitudinal consistency alongside divergent RSFC change patterns. Together, these findings provide a multilevel framework for characterizing heterogeneity within the psychosis risk state and help integrate dimensional and categorical perspectives on clinical and neural variation.

## Data Availability

The data used in this study were obtained from the Accelerating Medicines Partnership Schizophrenia (AMP SCZ) program and are available through the NIMH Data Archive (NDA).

## References

1. Hasmi L, Pries L-K, Have M ten, Graaf R de, Dorsselaer S van, Bak M, et al. (2021): What makes the psychosis ‘clinical high risk’ state risky: psychosis itself or the co-presence of a non-psychotic disorder? Epidemiol Psychiatr Sci 30: e53.

2. Catalan A, Salazar de Pablo G, Aymerich C, Damiani S, Sordi V, Radua J, et al. (2021): Neurocognitive Functioning in Individuals at Clinical High Risk for Psychosis. JAMA Psychiatry 78: 859–867.

3. Rutigliano G, Valmaggia L, Landi P, Frascarelli M, Cappucciati M, Sear V, et al. (2016): Persistence or recurrence of non-psychotic comorbid mental disorders associated with 6-year poor functional outcomes in patients at ultra high risk for psychosis. J Affect Disord 203: 101–110.

4. Fusar-Poli P, Cappucciati M, Borgwardt S, Woods SW, Addington J, Nelson B, et al. (2016): Heterogeneity of Psychosis Risk Within Individuals at Clinical High Risk: A Meta-analytical Stratification. JAMA Psychiatry 73: 113–120.

5. Baldwin H, Radua J, Antoniades M, Haas SS, Frangou S, Agartz I, et al. (2022): Neuroanatomical heterogeneity and homogeneity in individuals at clinical high risk for psychosis. Transl Psychiatry 12: 297.

6. Fusar-Poli P, De Micheli A, Signorini L, Baldwin H, Salazar de Pablo G, McGuire P (2020): Real-world long-term outcomes in individuals at clinical risk for psychosis: The case for extending duration of care. EClinicalMedicine 28: 100578.

7. Salazar de Pablo G, Besana F, Arienti V, Catalan A, Vaquerizo-Serrano J, Cabras A, et al. (2021): Longitudinal outcome of attenuated positive symptoms, negative symptoms, functioning and remission in people at clinical high risk for psychosis: a meta-analysis. EClinicalMedicine 36: 100909.

8. Ferrarelli F, Mathalon D (2020): The prodromal phase: time to broaden the scope beyond transition to psychosis? Schizophr Res 216: 5–6.

9. Simon AE, Velthorst E, Nieman DH, Linszen D, Umbricht D, de Haan L (2011): Ultra high-risk state for psychosis and non-transition: a systematic review. Schizophr Res 132: 8–17.

10. Salazar de Pablo G, Soardo L, Cabras A, Pereira J, Kaur S, Besana F, et al. (2022): Clinical outcomes in individuals at clinical high risk of psychosis who do not transition to psychosis: a meta-analysis. Epidemiol Psychiatr Sci 31: e9.

11. Fusar-Poli P (2017): The Clinical High-Risk State for Psychosis (CHR-P), Version II. Schizophr Bull 43: 44–47.

12. Healey KM, Penn DL, Perkins D, Woods SW, Keefe RSE, Addington J (2018): Latent Profile Analysis and Conversion to Psychosis: Characterizing Subgroups to Enhance Risk Prediction. Schizophr Bull 44: 286–296.

13. Catalan A, Radua J, McCutcheon R, Aymerich C, Pedruzo B, González-Torres MÁ, et al. (2022): Examining the variability of neurocognitive functioning in individuals at clinical high risk for psychosis: a meta-analysis. Transl Psychiatry 12: 198.

14. Fusar-Poli P, Salazar de Pablo G, Correll CU, Meyer-Lindenberg A, Millan MJ, Borgwardt S, et al. (2020): Prevention of Psychosis: Advances in Detection, Prognosis, and Intervention. JAMA Psychiatry 77: 755–765.

15. Del Fabro L, Schmidt A, Fortea L, Delvecchio G, D’Agostino A, Radua J, et al. (2021): Functional brain network dysfunctions in subjects at high-risk for psychosis: A meta-analysis of resting-state functional connectivity. Neurosci Biobehav Rev 128: 90–101.

16. Sasabayashi D, Takahashi T, Takayanagi Y, Nemoto K, Ueno M, Furuichi A, et al. (2023): Resting state hyperconnectivity of the default mode network in schizophrenia and clinical high-risk state for psychosis. Cereb Cortex 33: 8456–8464.

17. Kebets V, Holmes AJ, Orban C, Tang S, Li J, Sun N, et al. (2019): Somatosensory-Motor Dysconnectivity Spans Multiple Transdiagnostic Dimensions of Psychopathology. Biol Psychiatry 86: 779–791.

18. Wang HR, Liu Z-Q, Nomi JS, Schleifer CH, Bearden CE, Misic B, et al. (2026): Multivariate Resting-State Functional Connectivity Features Linked to Transdiagnostic Psychopathology in Early Psychosis. Biol Psychiatry 100: 178–188.

19. Wang HR, Liu Z-Q, Nakua H, Hegarty CE, Thies MB, Patel PK, et al. (2025): Decoding Early Psychoses: Unraveling Stable Microstructural Features Associated With Psychopathology Across Independent Cohorts. Biol Psychiatry 97: 167–177.

20. Nakua H, Yu J-C, Abdi H, Hawco C, Voineskos A, Hill S, et al. (2024): Comparing the stability and reproducibility of brain-behavior relationships found using canonical correlation analysis and partial least squares within the ABCD sample. Netw Neurosci 8: 576–596.

21. McIntosh AR, Lobaugh NJ (2004): Partial least squares analysis of neuroimaging data: applications and advances. NeuroImage 23 Suppl 1: S250–263.

22. Tang X, Wei Y, Pang J, Xu L, Cui H, Liu X, et al. (2025): Identifying neurobiological heterogeneity in clinical high-risk psychosis: a data-driven biotyping approach using resting-state functional connectivity. Schizophrenia 11: 13.

23. Sullivan PF, Geschwind DH (2019): Defining the Genetic, Genomic, Cellular, and Diagnostic Architectures of Psychiatric Disorders. Cell 177: 162–183.

24. Anderson KM, Collins MA, Chin R, Ge T, Rosenberg MD, Holmes AJ (2020): Transcriptional and imaging-genetic association of cortical interneurons, brain function, and schizophrenia risk. Nat Commun 11: 2889.

25. Fornito A, Arnatkevičiūtė A, Fulcher BD (2019): Bridging the Gap between Connectome and Transcriptome. Trends Cogn Sci 23: 34–50.

26. Arnatkeviciute A, Fulcher BD, Fornito A (2019): A practical guide to linking brain-wide gene expression and neuroimaging data. NeuroImage 189: 353–367.

27. Martins D, Giacomel A, Williams SCR, Turkheimer F, Dipasquale O, Veronese M (2021): Imaging transcriptomics: Convergent cellular, transcriptomic, and molecular neuroimaging signatures in the healthy adult human brain. Cell Rep 37: 110173.

28. Addington J, Liu L, Perkins DO, Carrion RE, Keefe RSE, Woods SW (2017): The Role of Cognition and Social Functioning as Predictors in the Transition to Psychosis for Youth With Attenuated Psychotic Symptoms. Schizophr Bull 43: 57–63.

29. Seidman LJ, Shapiro DI, Stone WS, Woodberry KA, Ronzio A, Cornblatt BA, et al. (2016): Association of Neurocognition With Transition to Psychosis: Baseline Functioning in the Second Phase of the North American Prodrome Longitudinal Study. JAMA Psychiatry 73: 1239–1248.

30. Fusar-Poli DP, Borgwardt DS, Bechdolf DA, Addington DJ, Riecher-Rössler DA, Schultze-Lutter DF, et al. (2013): The Psychosis High-Risk State: A Comprehensive State-of-the-Art Review. JAMA Psychiatry 70: 107.

31. Ma Q, Sahakian BJ, Zhang B, Li Z, Yu J-T, Li F, et al. (2025): Neural correlates of device-based sleep characteristics in adolescents. Cell Rep 44. 10.1016/j.celrep.2025.115565

32. Vieira S, Bolton T a. W, Schöttner M, Baecker L, Marquand A, Mechelli A, Hagmann P (2024): Multivariate brain-behaviour associations in psychiatric disorders. Transl Psychiatry 14: 231.

33. Drysdale AT, Grosenick L, Downar J, Dunlop K, Mansouri F, Meng Y, et al. (2017): Resting-state connectivity biomarkers define neurophysiological subtypes of depression. Nat Med 23: 28–38.

34. Wannan CMJ, Nelson B, Addington J, Allott K, Anticevic A, Arango C, et al. (2024): Accelerating Medicines Partnership® Schizophrenia (AMP® SCZ): Rationale and Study Design of the Largest Global Prospective Cohort Study of Clinical High Risk for Psychosis. Schizophr Bull 50: 496–512.

35. Esteban O, Markiewicz CJ, Blair RW, Moodie CA, Isik AI, Erramuzpe A, et al. (2019): fMRIPrep: a robust preprocessing pipeline for functional MRI. Nat Methods 16: 111–116.

36. Schaefer A, Kong R, Gordon EM, Laumann TO, Zuo X-N, Holmes AJ, et al. (2018): Local-Global Parcellation of the Human Cerebral Cortex from Intrinsic Functional Connectivity MRI. Cereb Cortex 28: 3095–3114.

37. Tian Y, Margulies DS, Breakspear M, Zalesky A (2020): Topographic organization of the human subcortex unveiled with functional connectivity gradients. Nat Neurosci 23: 1421–1432.

38. Diedrichsen J, Balsters JH, Flavell J, Cussans E, Ramnani N (2009): A probabilistic MR atlas of the human cerebellum. NeuroImage 46: 39–46.

39. Abdi H, Williams LJ (2013): Partial least squares methods: partial least squares correlation and partial least square regression. Methods Mol Biol 930: 549–579.

40. Wang Y, Genon S, Dong D, Zhou F, Li C, Yu D, et al. (2023): Covariance patterns between sleep health domains and distributed intrinsic functional connectivity. Nat Commun 14: 7133.

41. Umeh A, Kumar J, Francis ST, Liddle PF, Palaniyappan L (2020): Global fMRI signal at rest relates to symptom severity in schizophrenia. Schizophr Res 220: 281–282.

42. Wong CW, Olafsson V, Tal O, Liu TT (2013): The amplitude of the resting-state fMRI global signal is related to EEG vigilance measures. NeuroImage 83: 10.1016/j.neuroimage.2013.07.057.

43. Hawrylycz MJ, Lein ES, Guillozet-Bongaarts AL, Shen EH, Ng L, Miller JA, et al. (2012): An anatomically comprehensive atlas of the adult human brain transcriptome. Nature 489: 391–399.

44. Abdi H (2010): Partial least squares regression and projection on latent structure regression (PLS Regression). WIREs Comput Stat 2: 97–106.

45. Zhou Y, Zhou B, Pache L, Chang M, Khodabakhshi AH, Tanaseichuk O, et al. (2019): Metascape provides a biologist-oriented resource for the analysis of systems-level datasets. Nat Commun 10: 1523.

46. Ward Jr. JH (1963): Hierarchical Grouping to Optimize an Objective Function. J Am Stat Assoc 58: 236–244.

47. Dunlop K, Grosenick L, Downar J, Vila-Rodriguez F, Gunning FM, Daskalakis ZJ, et al. (2024): Dimensional and Categorical Solutions to Parsing Depression Heterogeneity in a Large Single-Site Sample. Biol Psychiatry 96: 422–434.

48. Caspi A, Houts RM, Belsky DW, Goldman-Mellor SJ, Harrington H, Israel S, et al. (2014): The p Factor: One General Psychopathology Factor in the Structure of Psychiatric Disorders? Clin Psychol Sci J Assoc Psychol Sci 2: 119–137.

49. Cowan HR, Williams TF, Mittal VA, Addington J, Bearden CE, Cadenhead KS, et al. (2024): The Complex Latent Structure of Attenuated Psychotic Symptoms: Hierarchical and Bifactor Models of SIPS Symptoms Replicated in Two Large Samples at Clinical High Risk for Psychosis. Schizophr Bull 50: 1295–1309.

50. Supekar K, Cai W, Krishnadas R, Palaniyappan L, Menon V (2019): Dysregulated Brain Dynamics in a Triple-Network Saliency Model of Schizophrenia and Its Relation to Psychosis. Biol Psychiatry 85: 60–69.

51. Dong D, Wang Y, Chang X, Luo C, Yao D (2018): Dysfunction of Large-Scale Brain Networks in Schizophrenia: A Meta-analysis of Resting-State Functional Connectivity. Schizophr Bull 44: 168–181.

52. Menon V (2011): Large-scale brain networks and psychopathology: a unifying triple network model. Trends Cogn Sci 15: 483–506.

53. O’Neill A, Mechelli A, Bhattacharyya S (2019): Dysconnectivity of Large-Scale Functional Networks in Early Psychosis: A Meta-analysis. Schizophr Bull 45: 579–590.

54. Zhang Y, Xu L, Hu Y, Wu J, Li C, Wang J, Yang Z (2019): Functional Connectivity Between Sensory-Motor Subnetworks Reflects the Duration of Untreated Psychosis and Predicts Treatment Outcome of First-Episode Drug-Naïve Schizophrenia. Biol Psychiatry Cogn Neurosci Neuroimaging 4: 697–705.

55. Keane BP, Abrham YT, Cole MW, Johnson BA, Hu B, Cocuzza CV (2025): Functional dysconnectivity of visual and somatomotor networks yields a simple and robust biomarker for psychosis. Mol Psychiatry 30: 1539–1547.

56. Cicero DC, Jonas KG, Li K, Perlman G, Kotov R (2019): Common Taxonomy of Traits and Symptoms: Linking Schizophrenia Symptoms, Schizotypy, and Normal Personality. Schizophr Bull 45: 1336–1348.

57. Cao H, Wei X, Hu N, Zhang W, Xiao Y, Zeng J, et al. (2022): Cerebello-Thalamo-Cortical Hyperconnectivity Classifies Patients and Predicts Long-Term Treatment Outcome in First-Episode Schizophrenia. Schizophr Bull 48: 505–513.

58. Cao H, Chén OY, Chung Y, Forsyth JK, McEwen SC, Gee DG, et al. (2018): Cerebello-thalamo-cortical hyperconnectivity as a state-independent functional neural signature for psychosis prediction and characterization. Nat Commun 9: 3836.

59. Lányi O, Zahemszky D, Wenning AS, Engh MA, Molnár Z, Horváth AA, et al. (2026): Cerebello-Thalamo-Cortical Dysconnectivity in Schizophrenia Spectrum Disorders: A Resting-State Functional Magnetic Resonance Imaging Meta-Analysis. Biol Psychiatry Cogn Neurosci Neuroimaging 11: 329–346.

60. Rotaru DC, Lewis DA, Gonzalez-Burgos G (2012): The role of glutamatergic inputs onto parvalbumin-positive interneurons: relevance for schizophrenia. Rev Neurosci 23: 97–109.

61. Uhlhaas PJ, Singer W (2010): Abnormal neural oscillations and synchrony in schizophrenia. Nat Rev Neurosci 11: 100–113.

62. Cao H, Cannon TD (2019): Cerebellar Dysfunction and Schizophrenia: From “Cognitive Dysmetria” to a Potential Therapeutic Target. Am J Psychiatry 176: 498–500.

63. Zhao K, Rhee SY (2023): Interpreting omics data with pathway enrichment analysis. Trends Genet 39: 308–319.

64. Trubetskoy V, Pardiñas AF, Qi T, Panagiotaropoulou G, Awasthi S, Bigdeli TB, et al. (2022): Mapping genomic loci implicates genes and synaptic biology in schizophrenia. Nature 604: 502–508.

65. Mısır E, Akay GG (2023): Synaptic dysfunction in schizophrenia. Synapse 77: e22276.

66. Owen MJ, Legge SE, Rees E, Walters JTR, O’Donovan MC (2023): Genomic findings in schizophrenia and their implications. Mol Psychiatry 28: 3638–3647.

67. Mallard TT, Grotzinger AD, Smoller JW (2023): Examining the shared etiology of psychopathology with genome-wide association studies. Physiol Rev 103: 1645–1665.

68. Czarny P, Bialek K, Ziolkowska S, Strycharz J, Sliwinski T (2020): DNA damage and repair in neuropsychiatric disorders. What do we know and what are the future perspectives? Mutagenesis 35: 79–106.

69. Yang Z, Li M, Hu X, Xiang B, Deng W, Wang Q, et al. (2017): Rare damaging variants in DNA repair and cell cycle pathways are associated with hippocampal and cognitive dysfunction: a combined genetic imaging study in first-episode treatment-naive patients with schizophrenia. Transl Psychiatry 7: e1028.

70. Panuccio A, Biondo D, Picerni E, Genovesi B, Laricchiuta D (2022): Trauma-Related Internalizing and Externalizing Behaviors in Adolescence: A Bridge between Psychoanalysis and Neuroscience. Adolescents 2: 413–423.

71. Ryan AT, Addington J, Bearden CE, Cadenhead KS, Cornblatt BA, Mathalon DH, et al. (2018): Latent class cluster analysis of symptom ratings identifies distinct subgroups within the clinical high risk for psychosis syndrome. Schizophr Res 197: 522–530.

72. Gupta T, Cowan HR, Strauss GP, Walker EF, Mittal VA (2021): Deconstructing Negative Symptoms in Individuals at Clinical High-Risk for Psychosis: Evidence for Volitional and Diminished Emotionality Subgroups That Predict Clinical Presentation and Functional Outcome. Schizophr Bull 47: 54–63.

73. Allswede DM, Addington J, Bearden CE, Cadenhead K, Cornblatt B, Mathalon D, et al. (2020): Characterizing covariant trajectories of individuals at clinical high-risk for psychosis across symptomatic and functional domains. Am J Psychiatry 177: 164–171.

74. Deng W, Chong B, Addington J, Bearden CE, Cadenhead KS, Cornblatt BA, et al. (2025): Beyond the Descriptive: A Comprehensive, Multidomain Validation of Symptom Trajectories for Individuals at Clinical High Risk for Psychosis. Biol Psychiatry Cogn Neurosci Neuroimaging 10: 195–202.

75. Carrión RE, Correll CU, Auther AM, Cornblatt BA (2017): A Severity-Based Clinical Staging Model for the Psychosis Prodrome: Longitudinal Findings From the New York Recognition and Prevention Program. Schizophr Bull 43: 64–74.

