## Supplementary Materials for "Latent Brain-Symptom Dimensions Characterize Heterogeneity in Clinical High Risk for Psychosis"

### Supplementary Methods

#### Supplementary Methods 1. MRI Acquisition and Preprocessing

Scanning platforms included Siemens Prisma (VE11/XA30), Skyra (VE11), and Vida (XA31), alongside GE MR750 (DV26) and Philips Achieva (v5.7.1) scanners. Resting-state functional MRI data were acquired across sites using harmonized multiband echo-planar imaging (EPI) sequences (TR = 900 ms, TE = 35 ms, flip angle = 52°, multiband factor = 6, 2.4 mm isotropic resolution, ~5 min acquisition, 333–343 volumes). Both anterior–posterior (AP) and posterior–anterior (PA) phase-encoding directions were collected. During scanning, participants were instructed to fixate on a cross. In addition, high-resolution T1-weighted structural images were acquired on 3T Siemens, GE, and Philips scanners using harmonized 3D MPRAGE/TFE protocols (TR = 2500 ms, TE ≈ 2–4 ms, TI = 1000 ms, flip angle = 8°, voxel size = 0.8 mm isotropic, 208 slices). Detailed parameters for each modality are described in Harms et al. (1).

**Supplementary Methods 2. Control and Reliability Analyses**

*Cross-validated PLS*

To assess the generalizability of the LCs identified by PLS, we performed 5-fold cross-validation. Specifically, all CHR participants were randomly divided into five approximately equal subsets. In each of the five folds, one subset (20% of participants) was designated as the test set, while the remaining four subsets (80% of participants) served as the training set.

Within each fold, we conducted PLS analysis on the training datasets ($X_{train}$ and $Y_{train}$) to derive the RSFC saliences ($V_{train}$) and behavioral saliences ($U_{train}$). Using these saliences, we calculated the corresponding RSFC composite scores ($L_{X\,train} = X_{train} \times V_{train}$) and behavioral composite scores ($L_{Y\,train} = Y_{train} \times U_{train}$).

For the testing datasets, RSFC and behavioral composite scores were obtained by projecting $X_{test}$ and $Y_{test}$ onto the saliences derived from the training data ($L_{X\,test} = X_{test} \times V_{train}$, $L_{Y\,test} = Y_{test} \times U_{train}$). Subsequently, we calculated Pearson's correlations between $L_{X\ test}$ and $L_{Y\ test}$ for LC1 and LC3, and assessed their statistical significance using 10,000 permutation tests.

*PCA on behavioral measures*

To assess the robustness of our components across different methods, we conducted principal component analysis (PCA) on the z-scored behavioral measures, consistent with the processing used before PLS. Subsequently, we computed Pearson's correlations between the first four principal components and the behavioral saliences of LC1 and LC3.

*Behavior normalization*

Several behavioral measures used in the PLS analysis were not normally distributed. To minimize potential impacts on the PLS results, we performed quantile normalization on all behavioral variables and repeated the PLS analysis, correlating RSFC with the normalized behavioral data.

Furthermore, the behavioral variable representing total anxiety showed significant skewness. To evaluate its influence, we conducted an additional PLS analysis after excluding this variable. In both follow-up analyses, we assessed robustness by correlating the brain (or behavioral) saliences from control analysis with those from the original PLS model.

*Global signal regression*

The global signal was estimated as the mean gray matter time series (2) and regressed from each regional time series using the Computational Brain Imaging Group (CBIG) toolbox (3). RSFC was then recomputed from the residualized time series using Pearson correlations. The PLS analysis was repeated using the resulting RSFC data, and robustness was evaluated by correlating the RSFC and clinical saliences with those from the original PLS model.

We further examined associations between global BOLD fluctuation amplitude, defined as the temporal standard deviation of the mean global BOLD time series (4), and the RSFC and clinical composite scores of LC1 and LC3 using partial correlations controlling for age, sex, education, and head motion. Differences in correlation strength were assessed using Fisher $r$ to $z$ transformation with participant level bootstrap resampling.

*Inclusion of positive symptoms*

Given the clinical relevance of positive symptoms, we repeated the PLS analysis among participants with available Positive Symptoms CAARMS Harmonized with SIPS (PSYCHS) data ($n$ = 397), adding positive symptom severity to the 34 clinical variables. RSFC saliences were correlated with those from the original PLS analysis, whereas clinical salience correlations were calculated across the 34 shared variables.

**Supplementary Methods 3. Gene expression data preprocessing**

The brain gene expression data were preprocessed using a standard pipeline (https://github.com/BMHLab/AHBAprocessing) (5). Microarray probes were reannotated using annotations from Arnatkeviciute et al. (5), and probes not reliably mapped to genes were excluded. Following reannotation, 45,812 probes corresponding to 20,232 genes were retained. Probes with low expression (not exceeding background noise in 50% of samples) were further removed, and for each gene, the probe with the highest differential stability across donors was selected (6), yielding a set of 15,633 probes corresponding to 15,633 genes.

T1-weighted MRI data from six donors were preprocessed using FreeSurfer to enable precise assignment of tissue samples to brain regions, and the D-K atlas was reconstructed in each AHBA donor brain. Tissue samples were assigned based on spatial proximity using a distance threshold, including samples within 2 mm of regional boundaries. A threshold was then applied to prevent sample assignment beyond a specified distance (7), with samples within 2 mm (Euclidean distance) of regional boundaries being included. Gene expression data were then normalized for each subject using a scaled robust sigmoid (8,9) to reduce inter-individual differences. Finally, expression values were averaged across samples within each region, yielding a region × 15,633 gene expression matrix.

#### Supplemental Results

*Control and Reliability Analyses*

Correlations between salience patterns from the control analyses and those from the original PLS analysis are summarized in Supplementary Table S5.

PCA was reapplied to the clinical variables to examine consistency across analytic approaches. The corresponding principal components correlated with the clinical salience patterns of LC1 ($r = 0.55$) and LC3 ($r = 0.78$).

The GSR analysis showed moderate correspondence with the original salience patterns for LC1 (RSFC, $r = 0.43$, $p < 0.001$; clinical, $r = 0.46$, $p = 0.0068$) and stronger correspondence for LC3 (RSFC, $r = 0.64$, $p < 0.001$; clinical, $r = 0.91$, $p < 0.001$). We further examined whether global BOLD fluctuation amplitude was differentially associated with the RSFC and clinical composite scores of LC1 and LC3. Global BOLD fluctuation amplitude was associated with the RSFC composite score for both LC1 ($r = 0.53$, $p < 0.001$) and LC3 ($r = -0.11$, $p = 0.005$), with a higher association for LC1 than LC3 (Δ Fisher $z = 0.70$, 95% CI, 0.58 to 0.81). For the clinical composite scores, the associations were weaker for LC1 ($r = 0.10$, $p = 0.006$) and LC3 ($r = 0.06$, $p = 0.089$), and did not differ between dimensions (Δ Fisher $z = 0.04$, 95% CI, −0.06 to 0.13).

**Supplementary Figure S1. Enrichment analysis of PLS1+ genes of LC1.** (A) The Metascape enrichment network shows intra- and inter-cluster similarities among enriched terms. Node size reflects the number of input genes, and node color indicates cluster membership. (B) Enrichment analysis of genes transcriptionally associated with RSFC loadings of LC1 (FDR, $q < 0.01$). Circle size indicates the number of genes associated with each pathway.

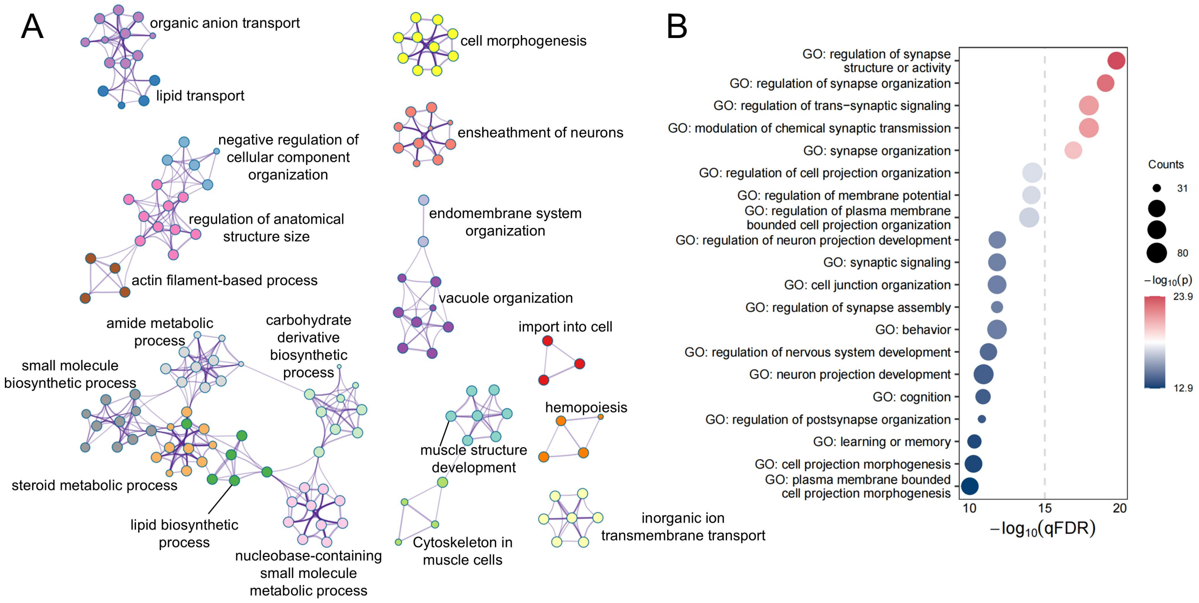

**Supplementary Figure S2. Transcriptomic decoding of regional RSFC loadings in LC3.** (A) Cortical distribution of regional PLS1 scores in the left hemisphere. (B) Association between RSFC loadings of LC3 and PLS1 scores ($r$ = 0.47, $p_{spin}$ = 0.0002). (C) Ranked PLS1 loading. (D) The Metascape enrichment network shows intra- and inter-cluster similarities among enriched terms. Node size reflects the number of input genes, and node color indicates cluster membership. (E) Enrichment analysis of genes transcriptionally associated with RSFC loadings of LC3 (FDR, $q$ < 0.01). Circle size indicates the number of genes associated with each pathway.

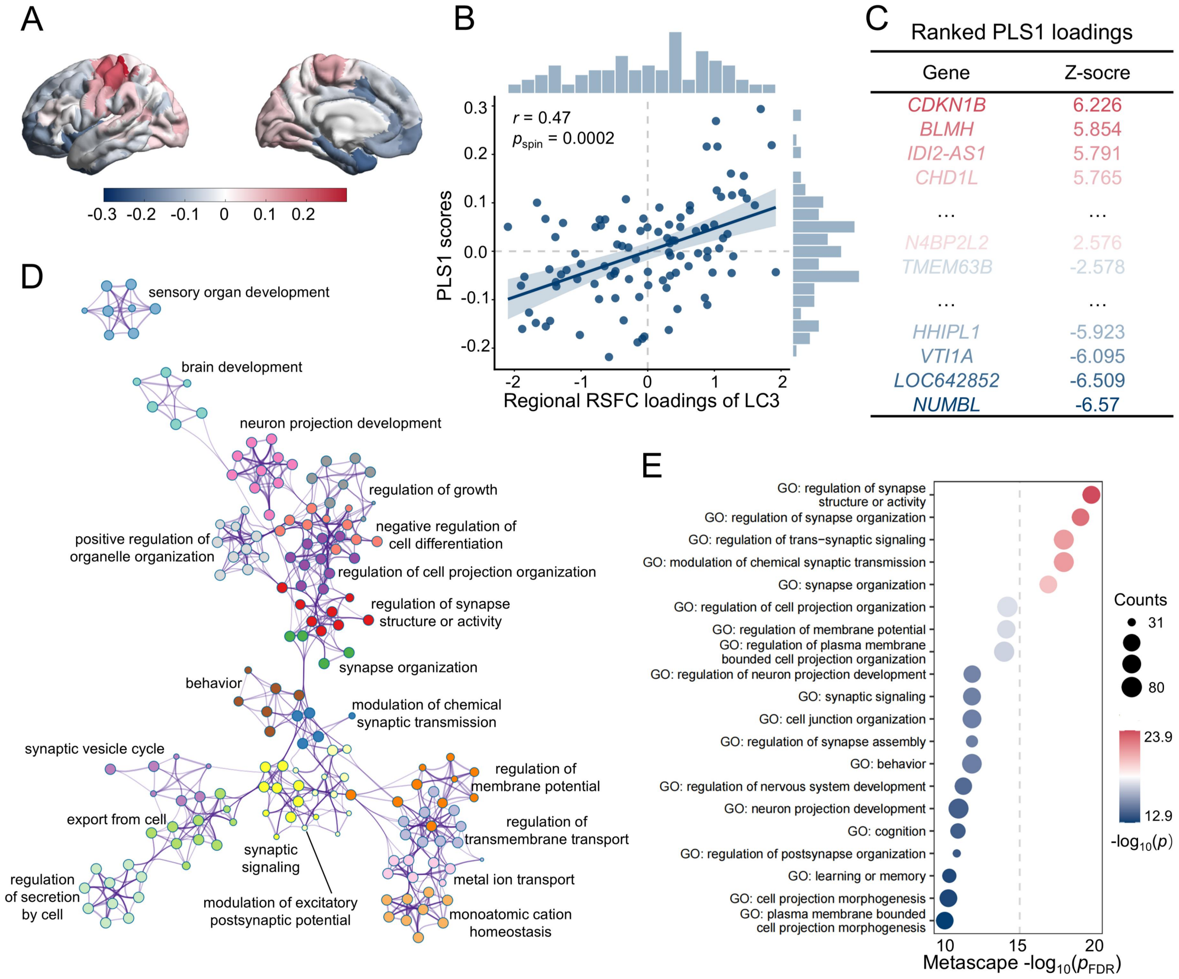

**Supplementary Figure S3. Enrichment analysis of PLS1+ genes of LC3.** (A) The Metascape enrichment network shows intra- and inter-cluster similarities among enriched terms. Node size reflects the number of input genes, and node color indicates cluster membership. (B) Enrichment analysis of genes transcriptionally associated with RSFC loadings of LC3 (FDR, $q < 0.01$). Circle size indicates the number of genes associated with each pathway.

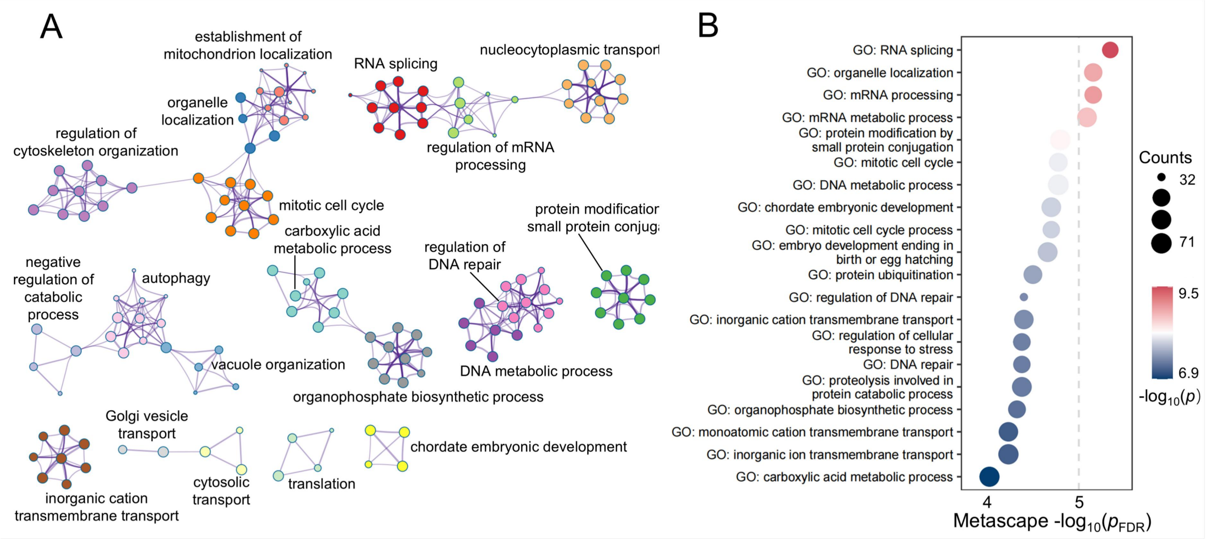

**Supplementary Figure S4. Evaluation of cluster number and clustering stability.** (A) Average silhouette index across 2 to 10 clusters. The highest silhouette index was observed for two clusters (0.510). (B) Calinski-Harabasz criterion across 2 to 10 clusters. The Calinski-Harabasz values were comparable for two and three clusters (700.230 and 700.784, respectively). Considering the higher silhouette index for two clusters and the comparable Calinski-Harabasz values for two and three clusters, two clusters were retained. (C) Null distribution obtained from permutation testing of the mean adjusted mutual information (AMI). The observed mean AMI was 0.455 and was significantly greater than expected by chance ($p = 0.001$), with the dashed line marking the observed mean AMI.

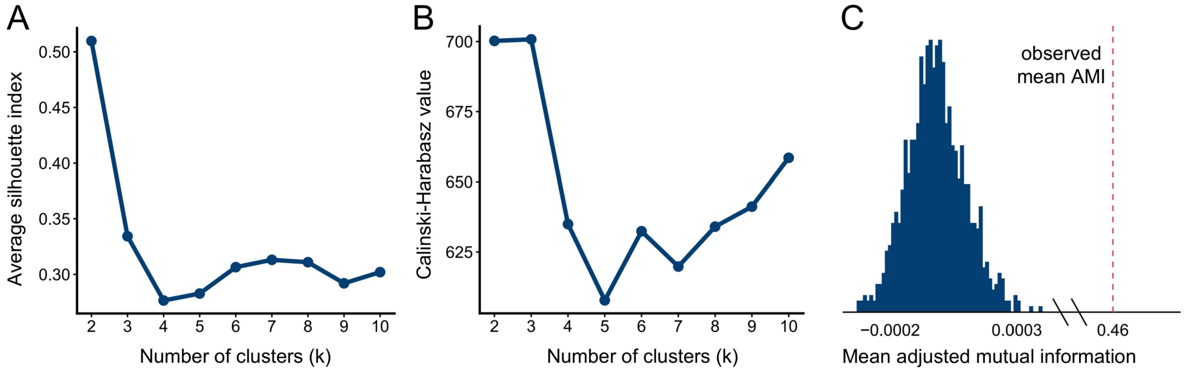

**Supplementary Figure S5. Longitudinal consistency of clinical differences across biotypes and healthy controls.** (A–C) Scatterplots show the correspondence between baseline and Month 2 *t*-values across clinical measures for Biotype 1 and Biotype 2, Biotype 1 and HC, and Biotype 2 and HC, respectively. Each point represents one clinical measure, and Pearson correlations quantify the similarity of group difference patterns between the two time points, indicating the extent to which baseline differences remained similar at Month 2. (D–F) *t*-values from pairwise comparisons of longitudinal change scores, calculated as Month 2 minus baseline, are shown for the corresponding pairwise group comparisons. * $p_{fdr} < 0.05$, ** $p_{fdr} < 0.01$, *** $p_{fdr} < 0.001$.

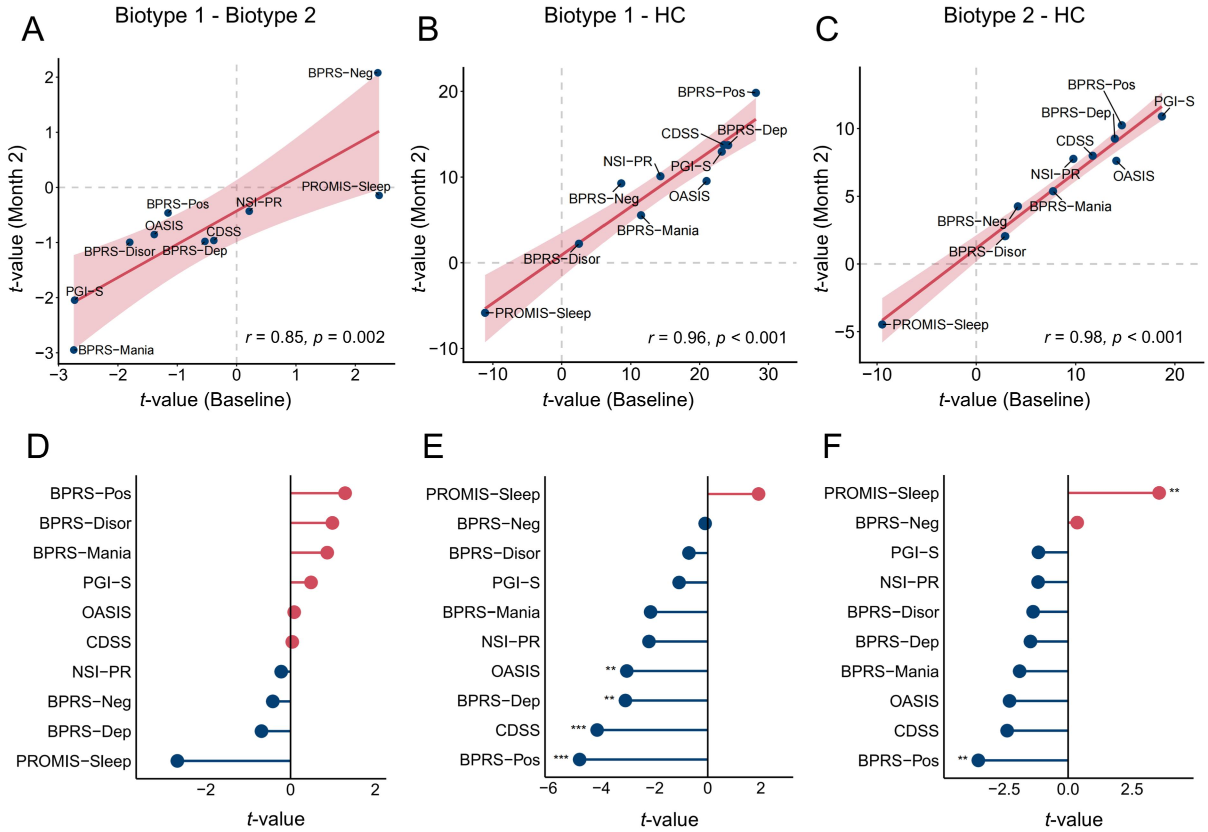

**Supplementary Figure S6. Longitudinal consistency of RSFC differences across biotypes and healthy controls.** Hexbin density plots show the correspondence between baseline and Month 2 *t*-value patterns across 34,716 RSFC connections. (A) Biotype 1 compared with Biotype 2, (B) Biotype 1 compared with HC, and (C) Biotype 2 compared with HC. Darker shading indicates a greater concentration of connections within the corresponding baseline and Month 2 *t*-value ranges. Positive correlations were observed for all three comparisons, supporting longitudinal consistency in the overall pattern of group differences.

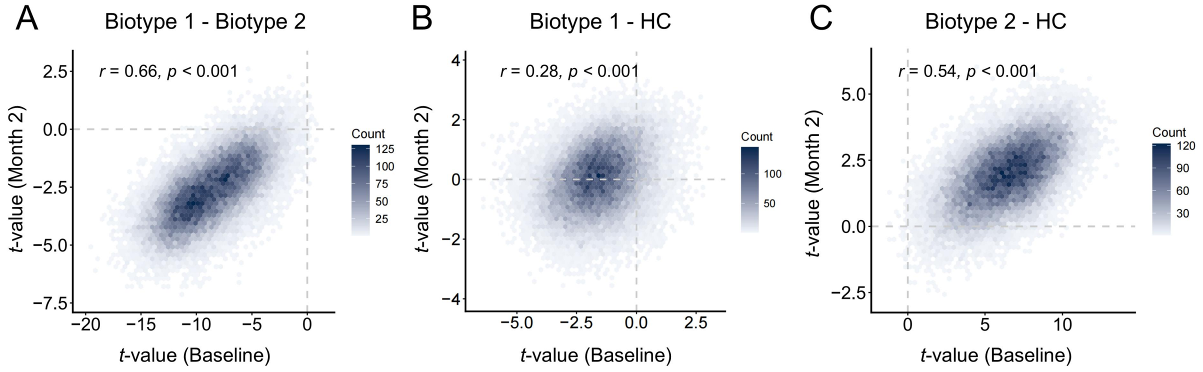

**Supplementary Figure S7. Group differences in RSFC at Month 2.** Edgewise RSFC differences at Month 2 were assessed using Welch's two-sample *t*-tests. (A) Biotype 1 compared with Biotype 2, (B) Biotype 1 compared with HC, and (C) Biotype 2 compared with HC. For each comparison, the upper panel shows the uncorrected t-value pattern, and the lower panel shows connections surviving Benjamini–Hochberg FDR correction ($q < 0.05$). At Month 2, Biotype 1 showed predominantly lower RSFC than Biotype 2, whereas Biotype 2 showed predominantly higher RSFC than HC. No connections between Biotype 1 and HC survived FDR correction.

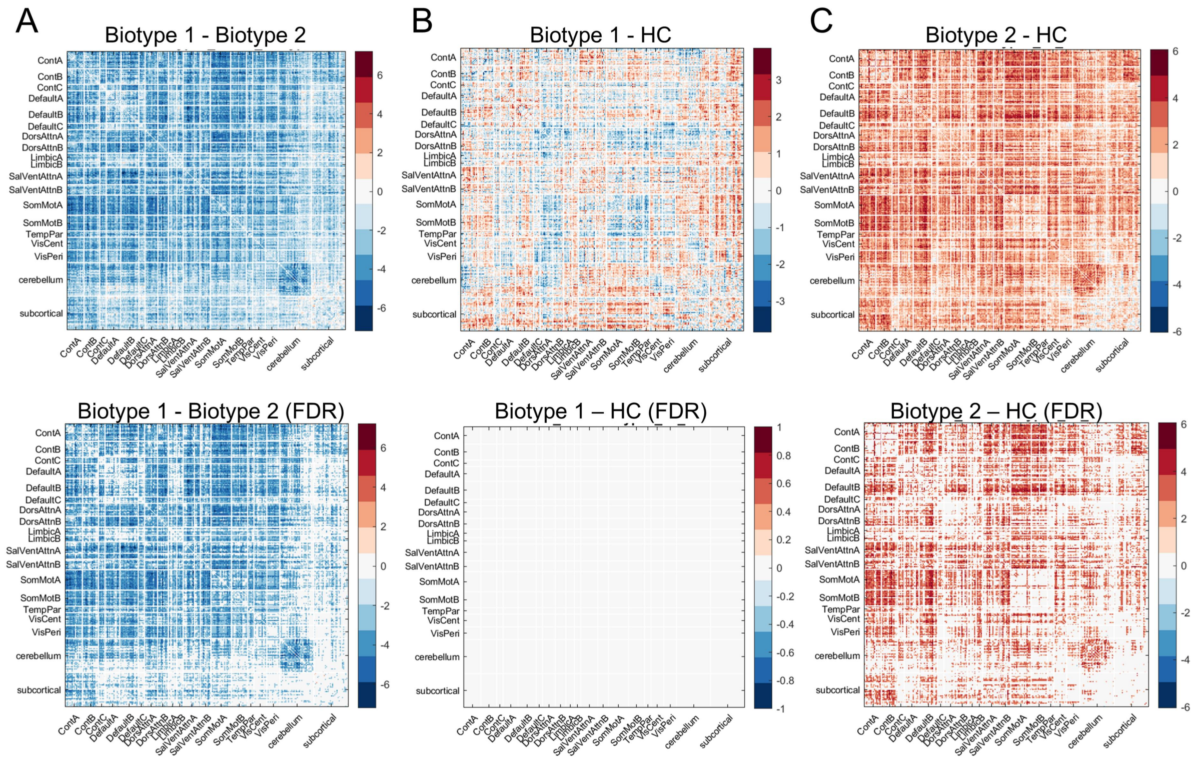

**Supplementary Figure S8. Group differences in longitudinal RSFC change.** Edgewise RSFC change scores were calculated as Month 2 minus baseline and compared between groups using Welch's two-sample *t*-tests. (A) Biotype 1 compared with Biotype 2, (B) Biotype 1 compared with HC, and (C) Biotype 2 compared with HC. For each comparison, the upper panel shows the uncorrected *t*-value pattern, and the lower panel shows connections surviving Benjamini–Hochberg FDR correction ($q < 0.05$). Biotype 1 showed predominantly higher change scores than Biotype 2. Compared with HC, no connections in Biotype 1 survived FDR correction, whereas Biotype 2 showed predominantly lower change scores.

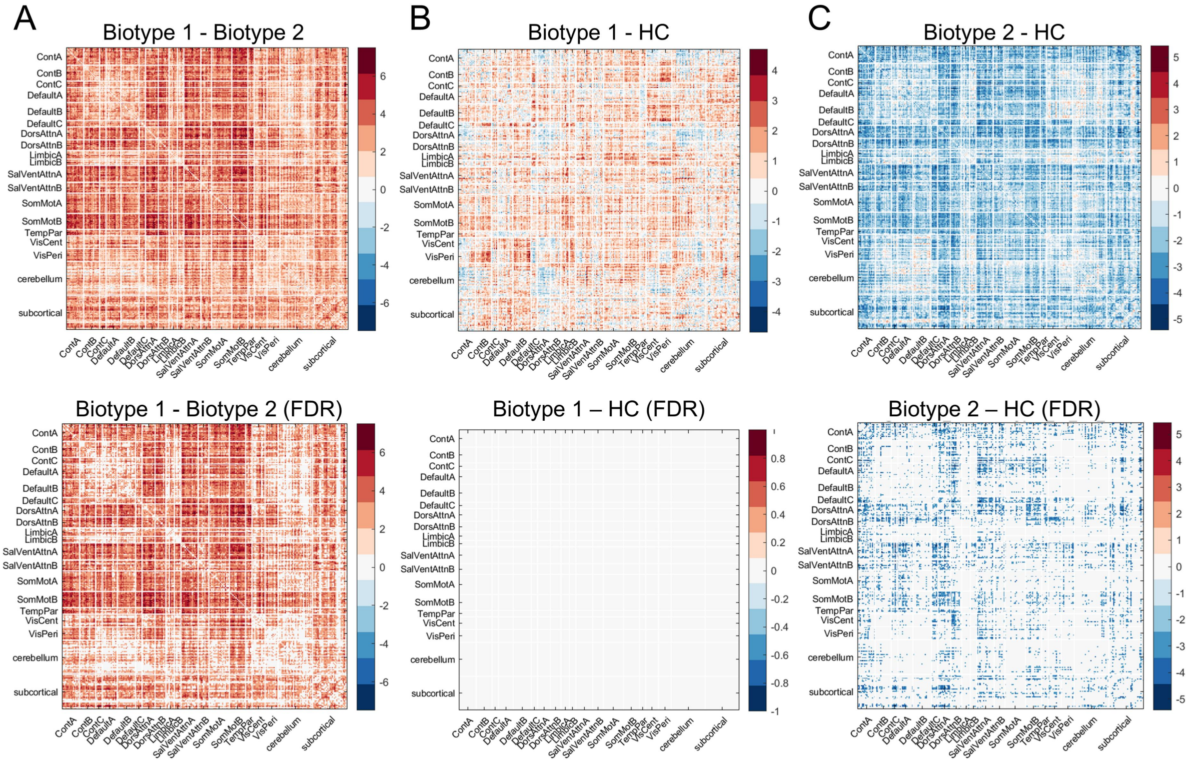

**Supplementary Table S1. Demographic and clinical characteristics of CHR participants included in the PLS analysis (*N* = 701) and healthy controls with available RSFC data (*N* = 244), including assessment instruments and variables.** Continuous variables are presented as mean (standard deviation), and categorical variables as n (%). For healthy controls, the mean and standard deviation for each measure were calculated using all available data. For comparisons between biotypes identified by hierarchical clustering, NSI-PR items were combined into a total score, and BPRS items were grouped into five subscales (i.e., positive symptoms, depression, negative symptoms, mania, and disorientation).

| **Measures** | | **Variable** | **CHR (*N* = 701)** | **HC ($N_{RSFC}$ = 244)** |
|---|---|---|---|---|
| Demographics | | Age, years | 21.64 (3.83) | 21.65 (3.51) |
| | | Female sex, *n* (%) | 433 (61.77%) | 145 (59.43%) |
| | | Education, years | 13.42 (2.72) | 14.05 (2.56) |
| Head Motion | | FD | 0.16 (0.06) | 0.15 (0.05) |
| Negative Symptom Inventory-Psychosis Risk (NSI-PR) (10) | | Motivation and pleasure | 1.82 (0.88) | 0.84 (0.73) |
| | | Diminished expression | 0.75 (0.92) | 0.28 (0.57) |
| | | Avolition | 1.70 (1.07) | 0.69 (0.73) |
| | | Asociality | 2.05 (1.05) | 0.98 (0.96) |
| | | Anhedonia | 1.60 (1.14) | 0.79 (0.85) |
| | | Blunted affect | 0.82 (1.02) | 0.29 (0.61) |
| Brief Psychiatric Rating Scale (BPRS) (11) | Positive symptoms | Hostility | 1.84 (1.01) | 1.28 (0.61) |
| | | Suspiciousness | 2.46 (1.26) | 1.09 (0.31) |
| | | Unusual thought content | 1.96 (1.14) | 1.04 (0.21) |
| | | Grandiosity | 1.38 (0.95) | 1.03 (0.21) |
| | | Hallucinations | 2.38 (1.38) | 1.03 (0.19) |
| | | Bizarre behavior | 1.15 (0.51) | 1.03 (0.23) |
| | Depression | Somatic concern | 2.06 (1.24) | 1.36 (0.75) |
| | | Anxiety | 3.48 (1.56) | 1.75 (1.01) |
| | | Depression | 3.10 (1.66) | 1.38 (0.80) |
| | | Guilt feelings | 2.41 (1.37) | 1.27 (0.64) |
| | | Suicidality | 1.73 (1.09) | 1.04 (0.24) |
| | Negative symptoms | Motor retardation | 1.19 (0.57) | 1.05 (0.26) |
| | | Blunted affect | 1.77 (1.04) | 1.30 (0.65) |
| | | Emotional withdrawal | 1.39 (0.77) | 1.11 (0.37) |
| | Mania | Excitement | 1.19 (0.60) | 1.03 (0.18) |
| | | Tension | 1.41 (0.77) | 1.07 (0.25) |
| | | Elevated mood | 1.26 (0.75) | 1.01 (0.15) |
| | | Distractibility | 1.48 (0.83) | 1.12 (0.40) |
| | | Motor hyperactivity | 1.27 (0.66) | 1.05 (0.31) |
| | Disorientation | Disorientation | 1.13 (0.49) | 1.08 (0.30) |
| | | Mannerisms & posturing | 1.04 (0.28) | 1.00 (0.00) |
| | Others | Self-neglect | 1.56 (0.92) | 1.09 (0.40) |
| | | Uncooperativeness | 1.09 (0.40) | 1.01 (0.11) |
| | | Conceptual disorganization | 1.27 (0.68) | 1.04 (0.27) |
| Calgary Depression Scale for Schizophrenia (CDSS) (12) | | Total score | 6.20 (4.55) | 1.02 (1.72) |
| The Patient Global Impression Scale-Severity (PGI-S) (13) | | PGI-S score | 2.68 (0.86) | 1.34 (0.64) |

| | | | |
|---|---|---|---|
| Overall Anxiety Severity and Impairment Scale (OASIS) (14) | Total score | 8.76 (4.00) | 3.09 (3.06) |
| 8-Item Patient-Reported Outcomes Measurement Information System (PROMIS) for Sleep (15) | Total score | 24.98 (6.30) | 30.22 (5.43) |

**Supplementary Table S2. Data availability and missingness for clinical measures included in the PLS analysis.** *N* indicates the number of CHR participants with available data for each individual clinical measure among the 838 participants who remained after MRI preprocessing quality control. Sample sizes therefore vary across measures.

| **Measures** | **Available *N*** | **Missing *N*** |
|---|---|---|
| Negative Symptom Inventory-Psychosis Risk (NSI-PR) | 751 | 87 |
| Brief Psychiatric Rating Scale (BPRS) | 759 | 79 |
| Calgary Depression Scale for Schizophrenia (CDSS) | 794 | 44 |
| The Patient Global Impression Scale-Severity (PGI-S) | 825 | 13 |
| Overall Anxiety Severity and Impairment Scale (OASIS) | 819 | 19 |
| 8-Item Patient-Reported Outcomes Measurement Information System (PROMIS) for Sleep | 817 | 21 |

**Supplementary Table S3. Demographic information of the six adult donors in the AHBA.**

| Characteristics | AHBA donors (*N* = 6) |
|---|---|
| Age, years, *M* (*SD*) | 42.50 (13.38) |
| Female sex, *n* (%) | 1.00 (16.67) |
| Hemispheric data availability | |
| Bilateral, *n* (%) | 2.00 (33.33) |
| Left hemisphere only, *n* (%) | 4.00 (66.67) |
| Postmortem interval (h), *M* (*SD*) | 20.67 (7.03) |

Abbreviation: AHBA, Allen Human Brain Atlas.

**Supplementary Table S4. Six assessment instruments used in the post hoc association analysis of the clinical dimensions.** Due to the use of available data, sample sizes varied across measures and are reported for each instrument.

| Instrument | Variable | *N* |
|---|---|---|
| Positive Symptoms CAARMS Harmonized with SIPS (16) | Total score | 394 |
| Columbia-Suicide Severity Rating Scale (C-SSRS) (17) | Suicidal intensity | 561 |
| Perceived Discrimination Scale (18) | Total score | 693 |
| Perceived Stress Scale (PSS) (19) | Total score | 690 |
| Social and Occupational Functioning Assessment Scale (SOFAS) (20) | Current total score | 599 |
| Global Functioning (21) | Current social scale total score | 698 |
| | Current role scale total score | 695 |
| Penn Computerized Neurocognitive Battery (PennCNB) | Short Penn list learning | 430 |
| | Motor praxis | 430 |
| | Short Penn continuous performance | 430 |
| | Penn emotion recognition | 430 |
| | Short fractal N-Back | 430 |
| | Digit symbol | 430 |
| | Short visual object learning | 430 |

**Supplementary Table S5. Absolute correlations between the RSFC (or behavioral) saliences obtained from the control analyses and those obtained from the original PLS analysis.**

| | Correlations with original RSFC saliences | | Correlations with original behavioral saliences | |
|---|---|---|---|---|
| | LC1 | LC3 | LC1 | LC3 |
| Behavior normalized | 0.99 | 0.98 | 0.99 | 0.98 |
| Skewed behaviors excluded | 1.00 | 1.00 | 1.00 | 0.99 |
| Confounds included | 0.96 | 1.00 | 0.80 | 0.98 |
| ComBat harmonization | 0.98 | 0.98 | 0.97 | 0.98 |
| Global signal regression | 0.43 | 0.64 | 0.46 | 0.91 |
| PSYCHS included | 0.67 | 0.67 | 0.77 | 0.72 |

#### References


1. Harms MP, Cho K-IK, Anticevic A, Bolo NR, Bouix S, Campbell D, *et al.* (2025): The MR neuroimaging protocol for the Accelerating Medicines Partnership® Schizophrenia Program. *Schizophrenia (Heidelb)* 11: 52.
2. Power JD, Plitt M, Laumann TO, Martin A (2017): Sources and implications of whole-brain fMRI signals in humans. *NeuroImage* 146: 609–625.
3. Kebets V, Holmes AJ, Orban C, Tang S, Li J, Sun N, *et al.* (2019): Somatosensory-Motor Dysconnectivity Spans Multiple Transdiagnostic Dimensions of Psychopathology. *Biological Psychiatry* 86: 779–791.
4. Wong CW, Olafsson V, Tal O, Liu TT (2013): The amplitude of the resting-state fMRI global signal is related to EEG vigilance measures. *Neuroimage* 83: 10.1016/j.neuroimage.2013.07.057.
5. Arnatkeviciute A, Fulcher BD, Fornito A (2019): A practical guide to linking brain-wide gene expression and neuroimaging data. *Neuroimage* 189: 353–367.
6. Hawrylycz M, Miller JA, Menon V, Feng D, Dolbeare T, Guillozet-Bongaarts AL, *et al.* (2015): Canonical genetic signatures of the adult human brain. *Nat Neurosci* 18: 1832–1844.
7. Romero-Garcia R, Whitaker KJ, Váša F, Seidlitz J, Shinn M, Fonagy P, *et al.* (2018): Structural covariance networks are coupled to expression of genes enriched in supragranular layers of the human cortex. *Neuroimage* 171: 256–267.
8. Fulcher BD, Fornito A (2016): A transcriptional signature of hub connectivity in the mouse connectome. *Proc Natl Acad Sci U S A* 113: 1435–1440.
9. Fulcher BD, Little MA, Jones NS (2013): Highly comparative time-series analysis: the empirical structure of time series and their methods. *J R Soc Interface* 10: 20130048.
10. Strauss GP, Walker EF, Pelletier-Baldelli A, Carter NT, Ellman LM, Schiffman J, *et al.* (2023): Development and Validation of the Negative Symptom Inventory-Psychosis Risk. *Schizophr Bull* 49: 1205–1216.
11. Overall J, Gorham D (1962): The Brief Psychiatric Rating-Scale. *Psychol Rep* 10: 799–812.
12. Addington D, Addington J, Maticka-Tyndale E (1993): Assessing depression in schizophrenia: the Calgary Depression Scale. *Br J Psychiatry Suppl* 39–44.
13. Yalcin I, Bump RC (2003): Validation of two global impression questionnaires for incontinence. *Am J Obstet Gynecol* 189: 98–101.
14. Norman SB, Cissell SH, Means-Christensen AJ, Stein MB (2006): Development and validation of an Overall Anxiety Severity And Impairment Scale (OASIS). *Depress Anxiety* 23: 245–249.
15. Yu L, Buysse DJ, Germain A, Moul DE, Stover A, Dodds NE, *et al.* (2011): Development of short forms from the PROMIS™ sleep disturbance and Sleep-Related Impairment item banks. *Behav Sleep Med* 10: 6–24.
16. Woods SW, Parker S, Kerr MJ, Walsh BC, Wijtenburg SA, Prunier N, *et al.* (2024): Development of the PSYCHS: Positive SYmptoms and Diagnostic Criteria for the CAARMS Harmonized with the SIPS. *Early Interv Psychiatry* 18: 255–272.
17. Posner K, Brown GK, Stanley B, Brent DA, Yershova KV, Oquendo MA, *et al.* (2011): The Columbia-Suicide Severity Rating Scale: initial validity and internal consistency findings from three multisite studies with adolescents and adults. *Am J Psychiatry* 168: 1266–1277.
18. Janssen I, Hanssen M, Bak M, Bijl RV, de Graaf R, Vollebergh W, *et al.* (2003): Discrimination and delusional ideation. *Br J Psychiatry* 182: 71–76.
19. Cohen S, Kamarck T, Mermelstein R (1983): A global measure of perceived stress. *J Health Soc Behav* 24: 385–396.
20. Goldman HH, Skodol AE, Lave TR (1992): Revising axis V for DSM-IV: a review of measures of social functioning. *Am J Psychiatry* 149: 1148–1156.
21. Cornblatt BA, Auther AM, Niendam T, Smith CW, Zinberg J, Bearden CE, Cannon TD (2007): Preliminary findings for two new measures of social and role functioning in the prodromal phase of schizophrenia. *Schizophr Bull* 33: 688–702.
